# The impact of observation models on epidemic inference for emerging infectious diseases: SARS-CoV-2 as a case study

**DOI:** 10.64898/2026.01.27.26344924

**Authors:** Matthieu Domenech de Cellès, Sarah C. Kramer

## Abstract

Parameter estimation is often necessary to inform transmission models of infectious diseases. This estimation requires choosing an observation model, defined here as the probability distribution linking model outputs to observed data. Although potentially consequential, this choice has received little attention in the literature. Here, we compared eight observation models, including common distributions such as the Poisson, binomial, negative binomial, and normal. Using Bayesian inference, we fit an SIR-like model to daily case reports during the first wave of COVID-19 in Belgium, Finland, Germany, Sweden, Switzerland, and the UK. We found considerable differences in the log-likelihoods of the observation models, spanning three orders of magnitude between the best and the worst. Compared with the best models, the binomial, Poisson, and normal models received no support due to their rigid variance structures. Additionally, the binomial and Poisson models produced overly narrow prediction and confidence intervals, especially for key parameters such as the basic reproduction number. The other five models—each with a free dispersion parameter scaling the variance to the mean—performed much better, with the negative binomial model ranking among the top models in all countries. We conclude that flexible observation models are essential for transmission models to accurately capture all sources of uncertainty.

## 1 Introduction

The COVID-19 pandemic, caused by SARS-CoV-2, underscored the usefulness of mathematical models of disease transmission as tools for understanding and controlling emerging pathogens. Early in the pandemic, these models provided critical information on the transmissibility of SARS-CoV-2 [1], including undocumented infections [2], age-related differences in susceptibility to infection [3], and the impacts of non-pharmaceutical interventions (NPIs) enacted by governments to mitigate the epidemic [4–6]. In addition to these scientific insights, transmission models were routinely used to forecast epidemic trajectories and healthcare demand under various scenarios, thereby informing policymaking (although this application of modeling amid the inevitable uncertainties of an unfolding epidemic requires caution [7,8]).

A critical aspect of effective transmission models is reliable parametrization. Clinical or epidemiological studies can quickly provide reliable information, enabling modelers to fix key epidemiological parameters such as the incubation and infectious periods, the generation time, and the serial interval [9]. However, other parameters may remain uncertain, either because external data are unavailable or because the available data cannot easily be translated into the model parameter of interest. For example, although contact tracing studies could, in theory, inform the basic reproduction number (*R_0_*) in a given transmission model, this is often impractical because *R_0_* depends on the model’s structure and assumptions [10]. A common method to overcome this obstacle is to estimate unknown parameters by fitting transmission models to epidemiological data, an exercise known as model estimation or calibration.

When fitted to data, any transmission model can be decomposed into two distinct sub-models serving different purposes. The first, called the process model, captures the scientific understanding of the system by defining the latent variables and mechanisms that produce the data. For epidemics of emerging pathogens that require rapid response with off-the-shelf models, process models are typically variants of the SIR model that mathematically describe how unobserved contacts between susceptible and infected individuals lead to new infections. The second, called the observation model (also known as the measurement model), describes how the outputs of the process model, such as predicted case counts, relate to data, such as observed case counts.

Broadly, observation models aim to capture the data-generating processes leading from the process model outputs to the observations. For infectious diseases, these processes represent the complexities of disease surveillance systems, typically characterized by reporting delays [11,12], under-reporting of cases (e.g., because of the presence of asymptomatic infections or limited testing capacity for symptomatic infections, both issues that were severe during the early COVID-19 pandemic [13]), imperfect sensitivity and specificity of diagnostic tests (e.g., due to test cross-reactivity with other pathogens [14]), or multiple data streams (e.g., case incidence and seroprevalence data [15]). Due to these complexities, observation models can be as structurally elaborate as process transmission models [16]. In addition to these components, observation models include a probability distribution that captures the stochasticity of the data-generating process and allows the formulation of a likelihood function.

Another important role of observation models is to compensate for model-data discrepancies caused by the process model formulation. Indeed, as process models are necessary simplifications of complex biological systems [17,18], they cannot be expected to reproduce the data exactly, even if the data were perfect. Hence, all process models are arguably misspecified to some extent, reflecting a mismatch between the data-generating mechanisms and their modeled versions. In practice, higher variability in the probability distribution of the observation model—or, for that matter, any stochastic component of the process model—can help account for the uncertainty arising from this misspecification [19].

These considerations suggest that observation models, although generally not of direct scientific interest, are nevertheless important, especially given the ever-increasing abundance and complexity of data available [20]. Arguably, therefore, “the measurement model is as much a part of the model as the latent process model,” [21] and careful attention should be paid to both the process and observation models for accurate statistical inference. Despite this importance, the choice of probability distributions for observation models has received little attention in the disease modeling literature, with few examples of systematic comparisons based on real-world or simulated data [15,22]. (Note that, to simplify terminology, we henceforth restrict our definition of the observation model to this probability distribution, although, as discussed above, observation models encompass other components of the data-generating process.) To address this gap, we aimed to compare the performance of various observation models by fitting transmission models to case-incidence data during the first wave of COVID-19 across six countries.

## 2 Methods

### 2.1 Study period & Study populations

We modeled the transmission dynamics of SARS-CoV-2 from February 2020 to June 30, 2020, in six countries: Belgium, Finland, Germany, Sweden, Switzerland, and the UK. We focused on this period, which corresponds to the first wave of COVID-19 in Europe, as a case study of fitting transmission models to limited and uncertain data during an emerging epidemic. We selected these countries because of the availability of seroprevalence studies, which provided estimates of under-reporting that informed the reporting probability in our model (Table 1). Following Ref. [4], we began model simulations 30 days before the first date when the cumulative number of COVID-19 deaths exceeded 10. This criterion helped reduce the impact of imported cases on the model fit at the beginning of the study period. The corresponding start date for model simulations ranged from 6 February in Germany to 22 February in Finland (Table 1). The reported case counts between the start date and 29 February were assumed unreliable (due to the influence of imported cases) and treated as missing values. Hence, in all countries, the model fit was based on the data observed from March 1 to June 30, 2020.

**Table 1:**
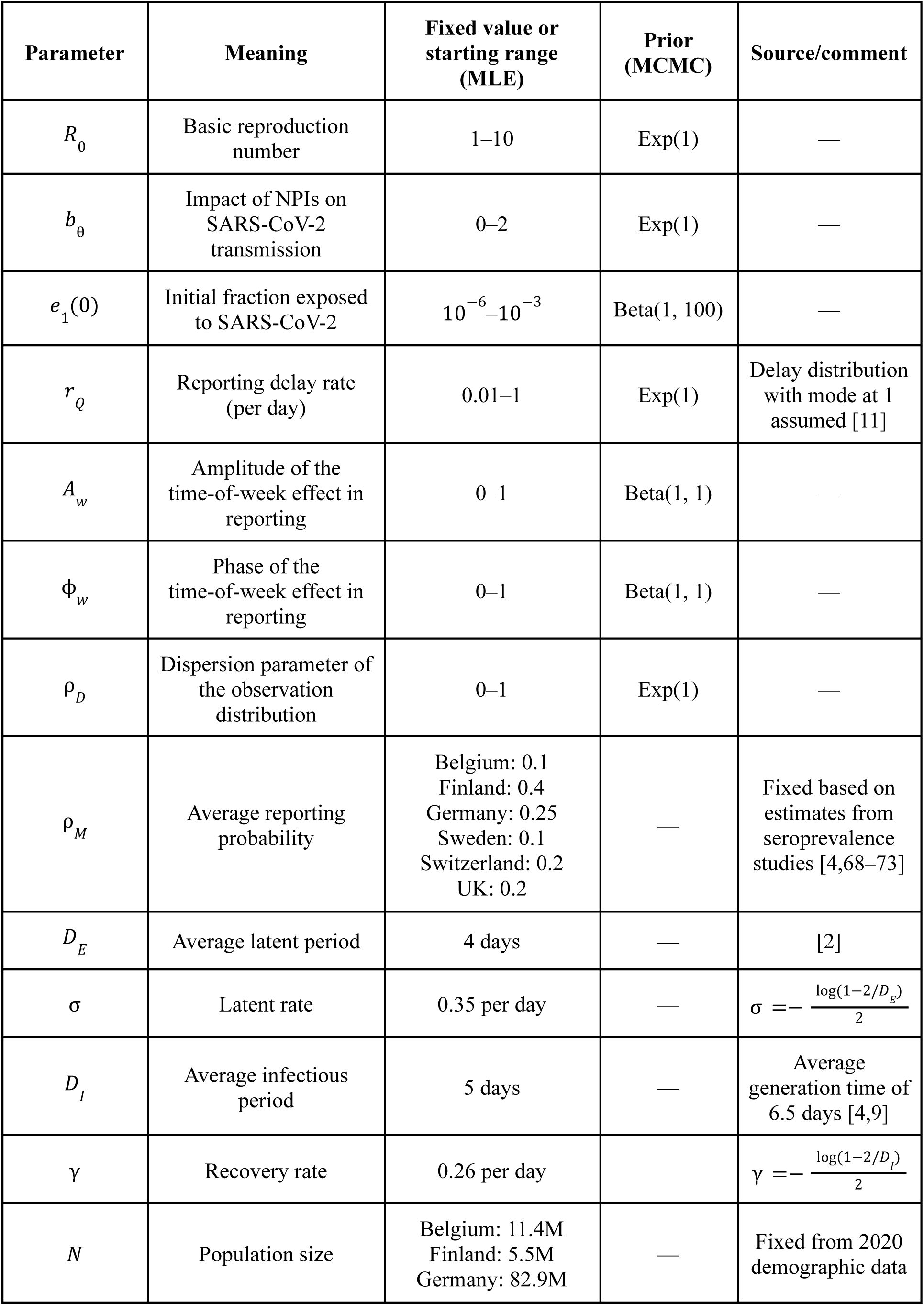
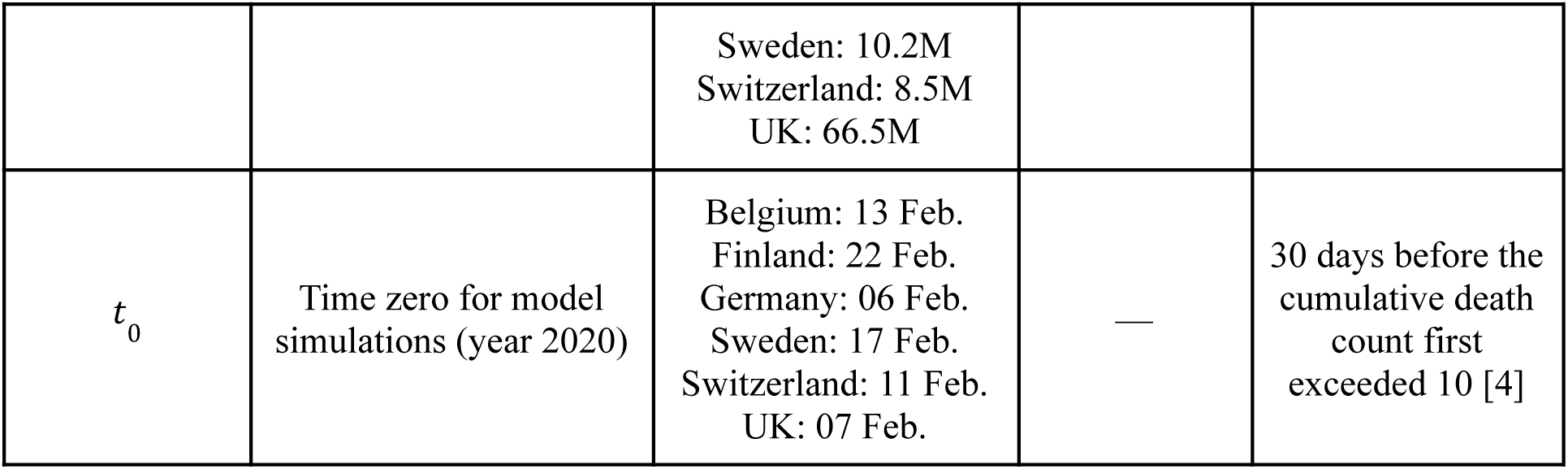
List of model parameters. MLE: Maximum Likelihood Estimation; MCMC: Markov Chain Monte Carlo estimation.

### 2.2 Data sources

The data consisted of the daily counts of COVID-19 confirmed cases (denoted by *O_t_*) and daily values of the so-called stringency index (denoted by θ*_t_*). As described elsewhere [23], this index is an aggregate measure that quantifies the strength of NPIs implemented by governments in response to the COVID-19 pandemic. The index is constructed systematically and ranges from 0 (no NPIs) to 100 (maximum number and strength of NPIs), enabling its use and comparison across countries. Importantly, however, this index does not directly quantify the impact of NPIs on SARS-CoV-2 transmission dynamics, as the same NPIs were implemented with varying effectiveness across countries [4]. All these data were available from the R package COVID19 [24,25] and are displayed in Fig. 1.

**Figure 1:**
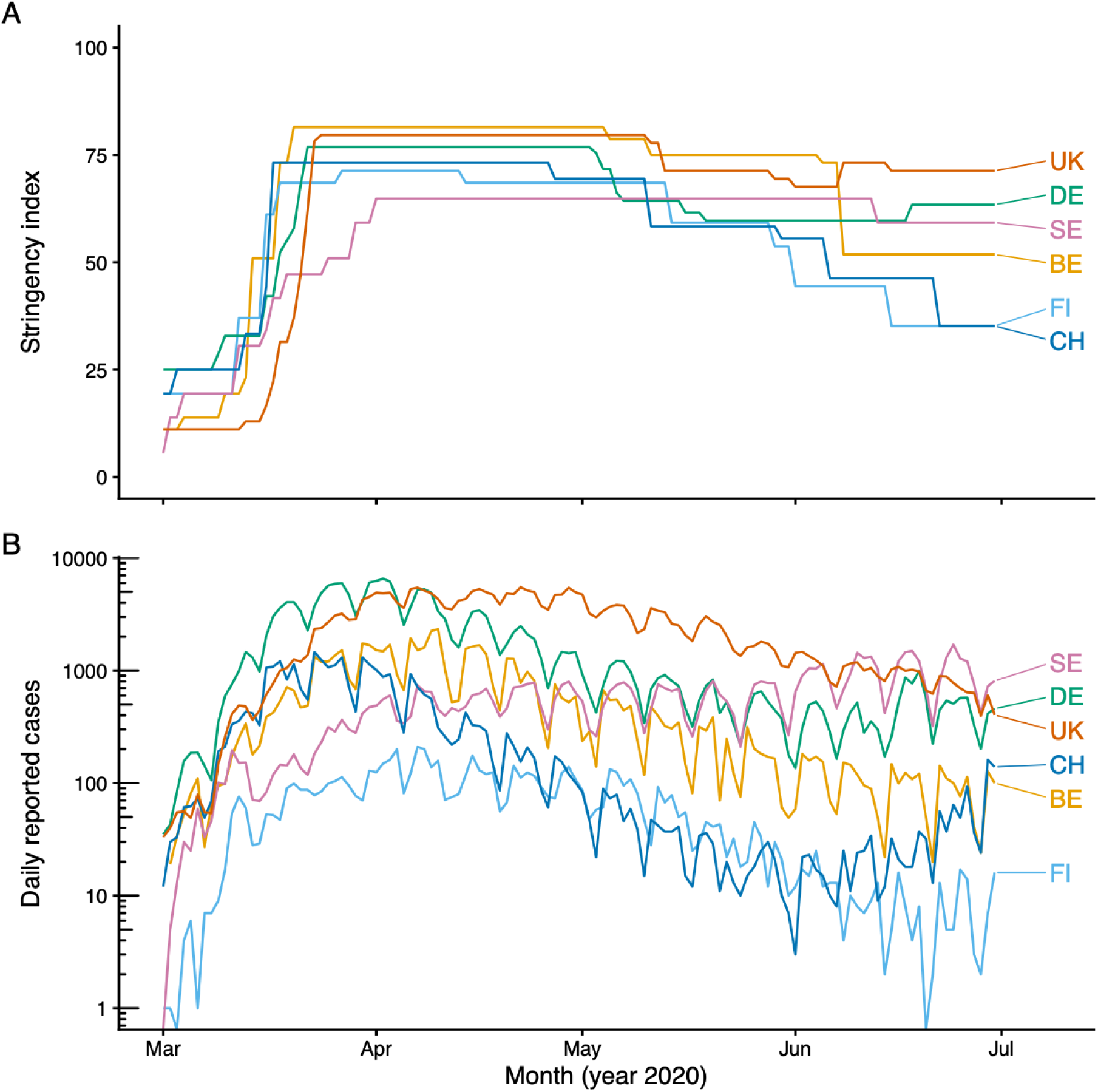
Stringency index and case incidence data in Belgium (BE), Finland (FI), Germany (DE), Sweden (SE), Switzerland (CH), and the UK. A: stringency index; B: daily incidence of reported cases. In panel B, the y-axis values are log_10_-transformed to better visualize the low values.

### 2.3 Process model

The process model was represented as a discrete-time SEIR model with a time step of 1 day to match the observation frequency. We deliberately ignored several complexities of SARS-CoV-2 epidemiology (e.g., age dependencies or other host heterogeneities such as variable susceptibility to infection, asymptomatic infections, or weather-related variations in transmission) to focus on an early epidemic situation in which, due to limited information, the simplest models are often the only practical choice.

The exposed and infected compartments were each divided into two sub-compartments to allow for a more realistic, negative binomial distribution of the latent and infectious periods [26,27]. Because of the time discretization, the average latent period equals 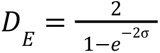 and the average infectious period is 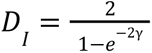 for this model (where σ represents the latency rate and γ the recovery rate) [28,29].

The model is represented schematically in Fig. 2 and specified by the following discrete-time map:

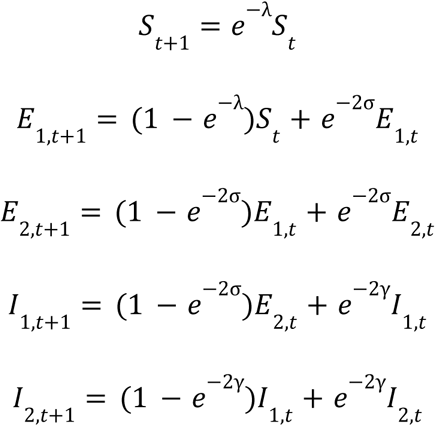

**Figure 2:**
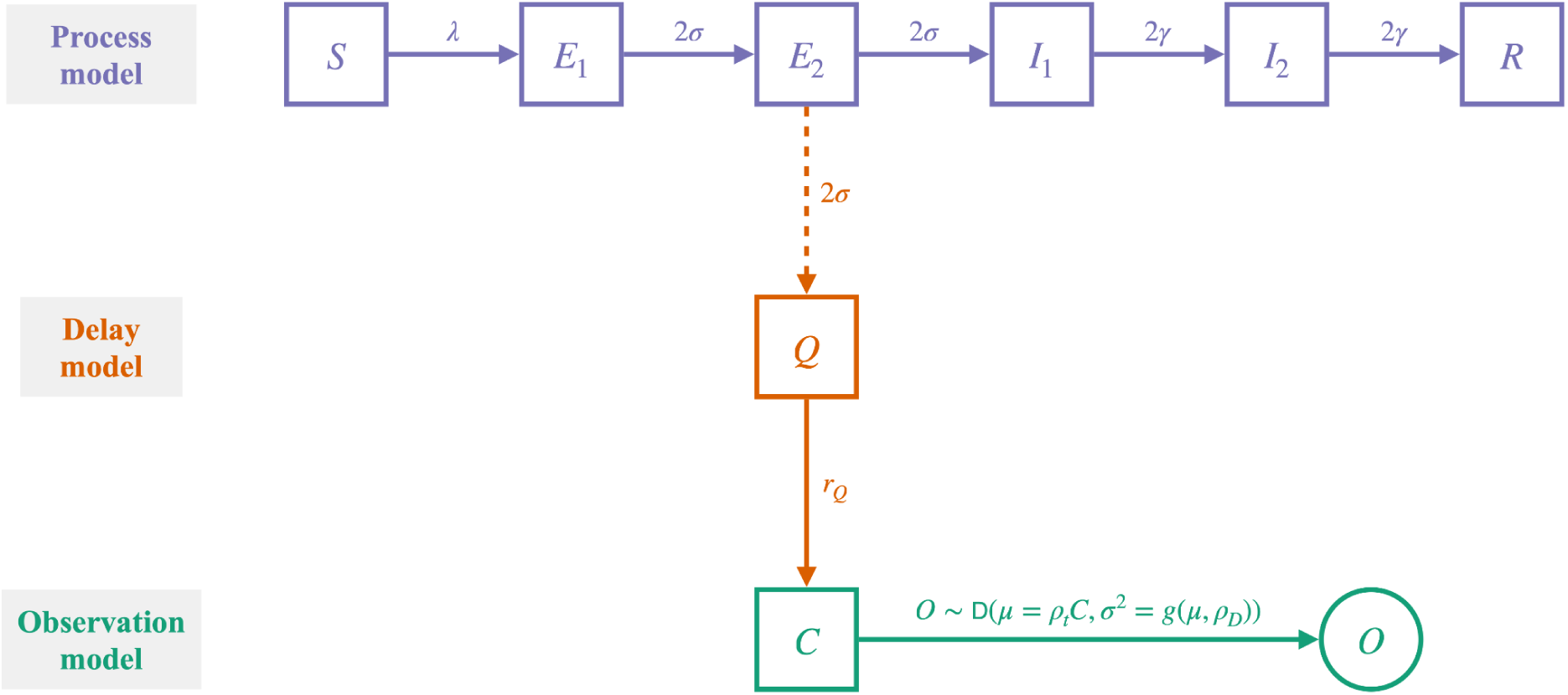
Model schematic. The observation model is here defined as the distribution D that maps the observed case count *O* to the total case count *C*, after accounting for delays (*Q* variable) and time-of-week effects in reporting. For all tested distributions, the mean (µ) is assumed to be identical, but the variance (σ^2^) differs in terms of its relationship with the mean (see Table 2 for the complete list of observation distributions and their corresponding variance structures). The dashed line indicates that the *E*_2_ → *Q* transition does not deplete the *E*_2_ compartment.

**Table 2:** Summary of observation distributions.

| Distribution | Parameters | Mean | Variance | Comment |
| --- | --- | --- | --- | --- |
| Poisson | $\lambda = \rho_t C_t$ | $\lambda = \rho_t C_t$ | $\lambda = \rho_t C_t$ | No dispersion parameter |
| Negative-binomial<br>(NegBinom) | $\mu = \rho_t C_t$<br>$k = 1/\rho_D$ | $\mu = \rho_t C_t$ | $\mu + \mu^2/k = \rho_t C_t + \rho_D(\rho_t C_t)^2$ | Tends to Poisson as<br>$\rho_D \rightarrow 0$ |
| Binomial<br>(Binom) | $n = C_t$<br>$p = \rho_t$ | $np = \rho_t C_t$ | $np(1 - p) = \rho_t C_t(1 - \rho_t)$ | No dispersion parameter<br>Finite support<br>($O_t \leq C_t$) |
| Beta-binomial<br>(BetaBinom) | $n = C_t$<br>$\alpha = \rho_t/\rho_D$<br>$\beta = (1 - \rho_t)/\rho_D$ | $n \frac{\alpha}{\alpha+\beta} = \rho_t C_t$ | $\frac{n\alpha\beta(\alpha+\beta+n)}{(\alpha+\beta)^2(\alpha+\beta+1)} = \rho_t C_t(1 - \rho_t) \frac{1+C_t\rho_D}{1+\rho_D}$ | Tends to Binomial as<br>$\rho_D \rightarrow 0$<br>Finite support<br>($O_t \leq C_t$) |
| Overdispersed<br>Binomial<br>(DispBinom) | $\mu = \rho_t C_t$<br>$\sigma = \sqrt{\mu(1 - \rho_t) + \rho_D \mu^2}$ | $\mu = \rho_t C_t$ | $\sigma^2 = \rho_t C_t(1 - \rho_t) + \rho_D(\rho_t C_t)^2$ | Overdispersed binomial<br>defined as a discretized<br>normal variable |
| Normal<br>(Norm) | $\mu = \rho_t C_t$<br>$\sigma = 10^3 \rho_D$ | $\mu = \rho_t C_t$ | $\sigma^2 = (10^3 \rho_D)^2$ | Equivalent to<br>least-squares estimation |
| Overdispersed<br>Student's $t$<br>(DispT) | $v = 4$ $\mu = \rho_t C_t$ $\sigma = 10^2 \rho_D \sqrt{\rho_t C_t}$ | $\mu = \rho_t C_t$ | $\sigma^2 \frac{v}{v-2} = 10^4 \rho_D^2 \rho_t C_t \frac{v}{v-2}$ | — |
| Log-normal<br>(logNorm) | $\mu = \log(\rho_t C_t) - \frac{\rho_D^2}{2}$ $\sigma = \rho_D$ | $e^{\mu + \sigma^2/2} = \rho_t C_t$ | $e^{2\mu + \sigma^2} (e^{\sigma^2} - 1) = (\rho_t C_t)^2 (e^{\rho_D^2} - 1)$ | — |

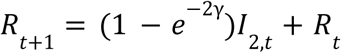

The force of infection was modeled as:

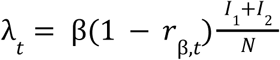

where β represents the mean transmission rate, related to the basic reproduction number through the equation: *R*_0_ = β*D*_1_. The variable *r*_β,*t*_ represents the time-varying (relative) reduction in SARS-CoV-2 transmission due to NPIs. Following previous work [30], we assumed a simple linear relationship between this variable and the stringency index:

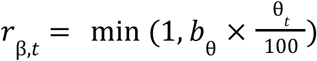

Here, the scaling parameter*b*_0_ > 0 quantifies the impact of NPIs on SARS-CoV-2 transmission. For this model, the effective reproduction number *R_e,t_* is:

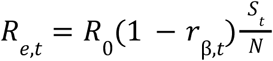

At time 0 (which varied between countries, see Table 1), we assumed that a fraction *e*_1_(0) of the population had been infected with SARS-CoV-2 (i.e., *E*_1_(0) = *N* × *e*_1_(0)) and that the rest of the population was susceptible to infection (*S*(0) = *N* × (1 – *e*_1_(0)). The other model compartments were initialized to 0.

### 2.4 Delay & Total Case Model

We defined the reporting delay as the time from the onset of a case’s infectiousness (exit from the *E*_2_ compartment) to the reporting of that case, conditional on reporting. To capture these delays, we defined a queuing variable *Q_t_*, modeled using the following equation:

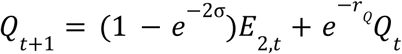

We then calculated the lagged, daily incidence rate *C_t_* (that is, the daily number of new infection cases) as follows:

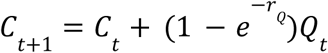

In this formulation, the parameter *r_Q_* represents the delay rate, and the delay distribution follows a Geometric distribution with support *n* ≥ 1 and mean 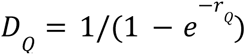. This choice was based on previous work showing that reporting delays generally had a highly skewed distribution with a mode near 0 [11].

### 2.5 Observation models

To complete the model’s specification, we defined a range of observation models that related the simulated total incidence rate *C_t_* to the observed incidence rate *O_t_*. Each observation model was represented by a probability distribution, with the same mean µ*_t_* = ρ*_t_C_t_* but a different mean-variance structure (Table 2). Here, ρ*_t_* represents the time-varying reporting probability, defined as:

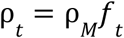

where ρ*_M_* is the average reporting probability. This parameter was fixed based on seroprevalence studies that estimated the under-reporting ratio by comparing reported cases with seroprevalence data during the first wave of COVID-19 (Table 1). Indeed, preliminary analyses indicated that this parameter was not identifiable from case incidence data due to a strong trade-off with the initial condition. This finding is consistent with earlier studies [31] and suggests that this parameter may be identified only with additional information from other data sources [15].

The variable *f_t_* represents a 1-week periodic function with unit mean, used to capture time-of-week variations in case reporting. These variations, apparent in Fig. 1B, are attributable to changes in testing effort over the week, e.g., reduced testing on weekends.

Following a modeling study in Germany [5], we used the following empirical function to model these variations:

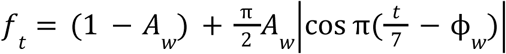

Here, time *t* is expressed in days, and the parameters *A_W_* and **φ***_W_* are the amplitude and the phase of the time-of-week effect in case reporting, both constrained to the 0–1 interval and estimated from the data. Equivalently, the weekday of peak case reporting is given by 7**φ***_W_*.

We considered eight different observation models, summarized in Table 2 and detailed below. Of those, four were discrete statistical distributions with probability mass functions (PMFs) *p_n_* defined over positive integers *n*: Poisson, negative binomial, binomial, and beta-binomial. The remaining four were continuous distributions with cumulative density functions (CDFs) *F*(*x*) over real numbers *x*: normal with constant variance, overdispersed binomial, log-normal, and Student’s *t* distribution. To ensure the comparability of all observation models, we discretized each continuous distribution as follows:

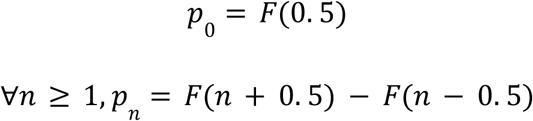

Although there are infinitely many possible discretizations, this one has been used in previous modeling studies [29].

#### 2.5.1 Poisson-based observation models

We first considered the Poisson distribution, a standard discrete model for case-count data that has sometimes been used in SARS-CoV-2 transmission models [32–34]. For this model, the variance equals the mean, so that the variance structure is fixed and lacks a free dispersion parameter.

Due to this limitation, a natural extension is the negative binomial model, another discrete model in which the variance scales with the square of the mean via an extra dispersion parameter (Table 2). This observation model is extremely common in the modeling literature on SARS-CoV-2 [28,30,35,36] and other pathogens, e.g., pertussis [37–39], measles [40], malaria [41,42], and Ebola [31].

#### 2.5.2 Binomial-based observation models

We considered three observation models based on the binomial distribution: the binomial distribution itself, the beta-binomial distribution, and an overdispersed binomial distribution. Like the Poisson distribution, the binomial distribution is a standard model for case count data, and its variance scales with the mean, with no extra dispersion parameter. In contrast, however, its support is limited to a finite range, such that *O_t_* ≤ *C_t_*.

An extension of this distribution is the beta-binomial distribution, a compound distribution where the probability of success is random and follows a Beta distribution. As a result, the variance structure is more flexible and controlled by a dispersion parameter that scales the variance with the square of the mean (Table 2). As for the Binomial distribution, the support of the beta-binomial is constrained to a finite range. See Ref. [22] for an example application of binomial and beta-binomial observation models for transmission models.

Finally, we tested another extension of the binomial distribution, initially proposed in Ref. [29] and used in subsequent transmission modeling studies [43–45]. In this model, a discretized normal distribution is used to augment the binomial’s variance via a dispersion parameter that scales the overall variance by the square of the mean (Table 2). Of note, unlike the binomial and beta-binomial distributions, the support of this distribution is not constrained.

#### 2.5.3 Normal-based observation models

We assessed three observation models related to the normal (or Gaussian) distribution: the normal, log-normal, and overdispersed Student’s *t* distributions.

For the normal distribution, we assumed a constant variance 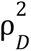. Due to the properties of the normal’s probability density function, the corresponding log-likelihood can be expressed as a sum of squared distances between the predicted and observed incidence rates. Hence, this observation model is equivalent to least-squares estimation, a standard method in nonlinear modeling, with some examples in transmission modeling [46,47].

The log-normal distribution (*X* ∼ *LN*(µ, σ)) is defined as a positive random variable that follows a normal distribution on the log-scale (log *X* ∼ *N*(µ, σ)). Although we could not find examples of this observation model in the disease modeling literature, we considered it because it offers a natural extension of the normal distribution to positive, right-skewed data. For this model, the variance is proportional to the square of the mean (Table 2).

Finally, we considered an overdispersed Student’s *t* distribution with four degrees of freedom and variance proportional to the mean (Table 2). This observation model was employed in a previous COVID-19 modeling study in Germany [5]; its variance structure is justified by earlier theoretical work on observation noise through random subsampling [48].

### 2.6 Estimation protocol

For every observation model and every country, six parameters were assumed unknown and estimated from the data: the basic reproduction number (*R*_0_ ≥ 0); the impact of NPIs on SARS-CoV-2 transmission (*b*_θ_ ≥ 0); the rate of delay in case reporting (*r_Q_* ≥ 0); the initial fraction exposed to SARS-CoV-2 (*e*_1_ (0) ∈ [0, 1]); the amplitude of the time-of-week effect in case reporting (*A_W_* ∈ [0, 1]); and the phase of the time-of-week effect in case reporting (**φ***_W_* ∈ [0, 1]).

For all the observation models except the Poisson and Binomial models (which have no free dispersion parameter), the dispersion parameter (ρ*_D_* ≥ 0) was also estimated from the data. Because the effect of this parameter varied across the observation models, we used proportionality constants to approximately rescale it to the 0–1 interval (Table 2). This rescaling allowed us to use the same prior distribution for Bayesian estimation of this parameter (see below). We stress, however, that this parameter cannot be compared across observation models.

The estimation was conducted in two steps. First, we used trajectory matching to maximize the log-likelihood and locate the maximum likelihood estimates (MLEs). All estimated parameters were first transformed (using log or logit functions) to lie on the real line. The search was initialized from 1,000 starting parameter sets generated using a Latin hypercube design [49] over broad parameter ranges (Table 1). Subsequently, multiple rounds of estimation were performed, with the starting parameter sets for each new round narrowed based on the best parameter sets from the previous round.

Second, we used Markov Chain Monte Carlo (MCMC) to estimate the model parameters in a Bayesian setting. All parameters were estimated on the natural scale (no parameter transformation) and assigned weakly informative priors with large variance, either Exponential (for positive parameters) or Beta (for parameters in the interval 0–1) distributions (Table 1). We used a differential evolution MCMC algorithm [50], with five chains run in parallel. Each chain had 20,000–50,000 iterations and was initialized at the MLEs. The initial parameter matrix for generating differential evolution proposals was seeded with all parameter sets within the multivariate 99% confidence interval from the MLE estimation round. Convergence to the posterior distribution was assessed by visual inspection of the chains and by calculation of Gelman-Rubin diagnostic statistics [51,52], with values below 1.1 considered indicative of convergence. The last 10,000 iterations were treated as a sample of the posterior distribution.

### 2.7 Comparison of the observation models’ performance

To compare the observation models, we calculated the widely applicable information criterion (WAIC [53]) and the deviance information criterion (DIC [54]). Both criteria estimate out-of-sample predictive accuracy by balancing goodness-of-fit against a penalty for model complexity (the so-called effective number of parameters, denoted by p_WAIC_ for the WAIC); however, they differ in that the DIC evaluates the log-likelihood at a single point estimate, whereas the WAIC averages the log pointwise predictive density over the entire posterior distribution. Unlike the DIC, the WAIC also provides a direct estimate of its standard error via the pointwise (per-observation) log-predictive density contributions, enabling uncertainty quantification in model comparisons. Hence, the WAIC is generally preferred to the DIC [55]. We used the WAIC where applicable, but switched to the DIC due to the known instability of p_WAIC_ under model mispecification [56].

### 2.8 Numerical implementation

The transmission models were implemented using the POMP package (version 6.3) [57,58], which operates within R (version 4.4.1) [59]. Trajectory matching was performed using the “Sbplx” algorithm (based on Subplex [19]) available in the nloptr package [60]. The MCMC estimation was carried out with the BayesianTools package (using the “DEzs” sampler) [61]; convergence was diagnosed using tools from the MCMCvis package [62], and the WAIC was calculated using the loo package [63].

## 3 Results

### 3.1 Comparative performance of observation models

We first evaluated the performance of the different observation models by calculating WAIC differences (ΔWAIC, Fig. 3 and Tables S1–S6) and DIC differences (ΔDIC, Tables S1–S6) relative to the best model in each country. The Poisson and binomial observation models consistently ranked lowest in all countries, with ΔDIC values exceeding 700 (Tables S1–S6). The poor performance of these models was confirmed by the instability of the WAIC, with unrealistically high p_WAIC_ values indicating model misspecification (Tables S1–S6). Hence, neither of these two models received any support from the data.

**Figure 3:**
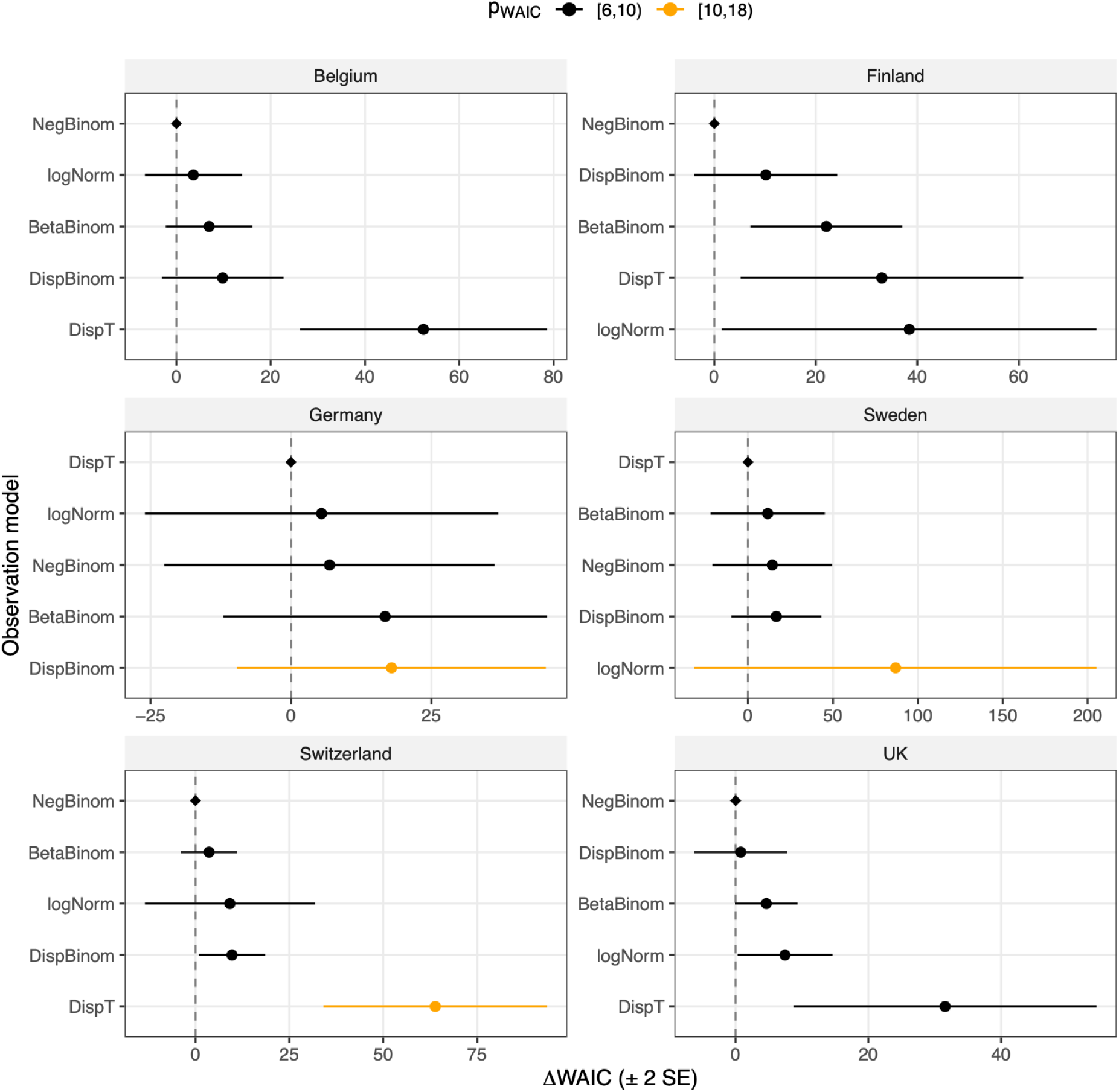
Comparative performance of the observation models. Abbreviations: WAIC, widely applicable information criterion; ΔWAIC, difference between the model’s WAIC and the best model with the lowest WAIC in that country (hence, ΔWAIC=0 for the best model); SE: standard error; p_WAIC_: effective number of parameters (to be compared to the nominal number of estimated parameters, which was seven for all the models presented here). NB: for visual clarity, the normal, Poisson, and binomial models are omitted from the figure, as these models received little support from the data.

Although better supported by the data, the normal model was consistently outperformed by the best model (ΔWAIC range: 43–334), with a rank intermediate between the Poisson and binomial models and the five overdispersed models. The only exception was Sweden, where the normal model ranked fourth (based on WAIC point estimates and DIC, Table S4) and performed better than the log-normal model.

Compared with the binomial, Poisson, and normal observation models, the five overdispersed variants received substantially more statistical support. Although the ΔWAIC point estimates suggested large performance differences, no single model performed best when ΔWAIC uncertainty was taken into account. Across countries, the number of best-performing models ranged from 2 (Finland) to 6 (Germany and Sweden). Across models, the negative binomial ranked among the best in all countries (and first in four countries); the overdispersed binomial and beta-binomial in five countries; the log-normal model in three countries; and the overdispersed *t* model in two countries. Interestingly, the latter model ranked first (according to WAIC point estimates) in Germany and Sweden but received little support in the other four countries.

### 3.2 Models’ fit to case incidence data

To visually compare the models’ performance, we ran simulations to assess model fit in each country. These simulations involved random sampling from the observation models and deterministic simulations of the process models at the Maximum A Posteriori (MAP) estimates, allowing us to assess the effect of the observation model on model fit. The results are presented in Fig. 4 for the UK and in Figs. S1–S5 for the other countries.

**Figure 4:**
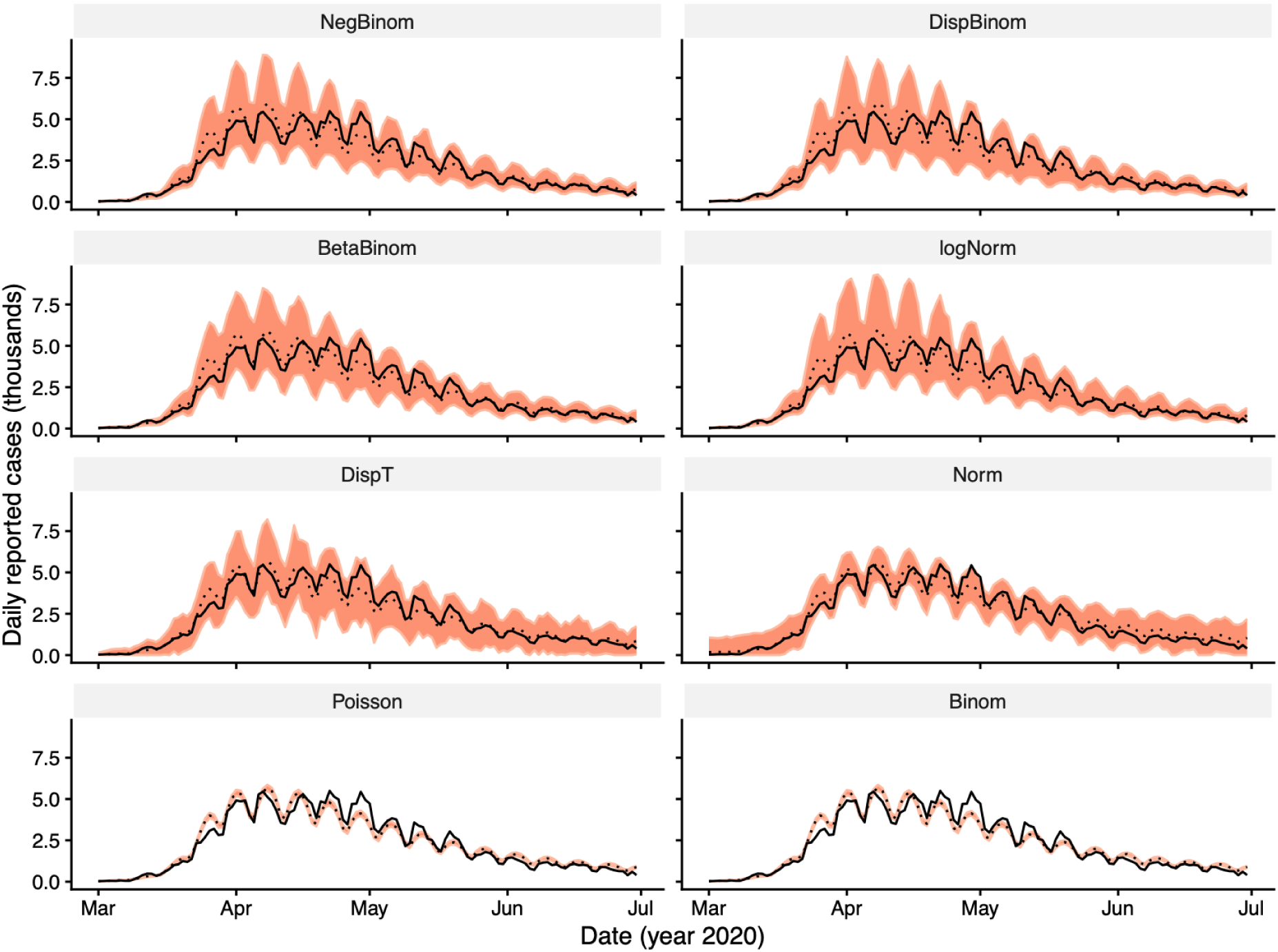
Models fit in the UK. The dotted line (ribbon) shows the mean (99% prediction interval) from 1,000 draws of the observation model, with the deterministic process model simulated at the maximum a posteriori estimate of the MCMC chain. Therefore, the predictive variability is only due to the observation model. The solid black lines represent the data.

Although the model fit was relatively accurate across all observation models in each country, differences in the variance structure were evident and could explain why some models performed worse. This was especially true for the Poisson and binomial models, which failed to capture the weekly peaks in reported cases because of their inflexible variance structures, which lack an extra dispersion parameter to adjust the variance. Although the normal model performed better, it was also hindered by this issue due to its fixed variance.

In contrast, the other five observation models fit the data much better. Their flexible variance structure—incorporating a dispersion parameter that controls how the variance scales with the mean—enabled them to capture systematic discrepancies between the model and the data, thereby reducing their impact on the model’s likelihood. As a result, these models could better adjust for model misspecification, which—despite a good overall fit—was particularly evident at the start of the study period in Finland and the UK and at the end in Sweden.

To complement these posterior predictive checks, we compared the pointwise log-likelihoods of the five overdispersed observation models over time (Fig. S6). At intermediate observed case counts, the models yielded very similar log-likelihoods in each country. Systematic differences appeared mainly at the extremes: in Finland and Sweden, the log-normal model assigned markedly lower likelihood to very small counts, while the *t* model assigned somewhat lower likelihood than the negative binomial at large counts in Belgium, Switzerland, and the UK, consistent with the WAIC-based ranking of these models. Overall, these results suggest that performance differences among these observation models arose from modest, cumulative discrepancies over many days concentrated in the tails of the incidence distribution, rather than from a small number of extreme outliers.

### 3.3 Impact of observation model on estimated model parameters

Next, we examined the parameter estimates and their variation across observation models (Fig. 5 and Tables S1–S6). Overall, the estimates were similar for the parameters capturing the time-of-week effect (*A_w_* and **φ***_w_*) and delay in case reporting (*r_Q_*), and the impact of NPIs on SARS-CoV-2 transmission (*b*_θ_). Regardless of the observation model, the results showed a substantial impact of NPIs, with some differences across countries (*b*_θ_ range for the best model: 0.96 to 1.40; Tables S1–S6). At the peak intensity of NPIs (i.e., the maximal stringency index), these estimates corresponded to a 60% reduction in transmission in Sweden, 90% in Belgium, Germany, and the UK, 95% in Switzerland, and ∼100% in Finland. Similarly, the time-of-week effect was fairly consistent across countries, with a common dip in case reporting on Sundays, although the effect’s amplitude varied (Fig. S7). There was also evidence of substantial reporting delays in all countries except Sweden, with medians ranging from 10 days in Germany to 22 days in the UK (Fig. S8). In Sweden, the reporting delays were less pronounced (median: 1 day; mean: 3 days).

**Figure 5:**
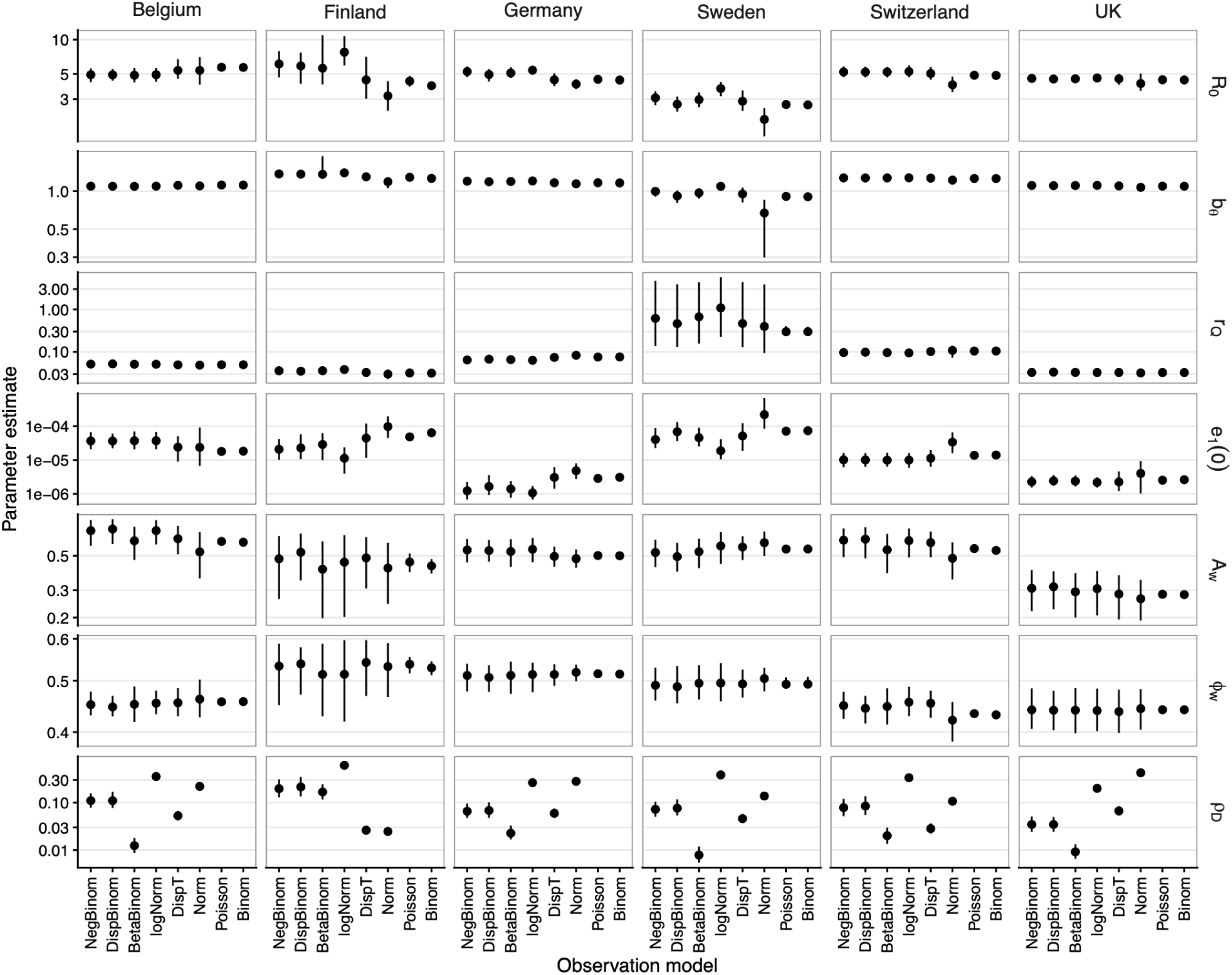
Parameter estimates across observation models and countries. The points and intervals represent the median and 99% credible intervals, respectively, calculated from 1,000 MCMC draws of the posterior distribution. Parameters: *R*_0_, basic reproduction number; *b*_θ_, impact of control measures; *r_Q_*, reporting delay rate (per day); *e*_1_ (0), initial fraction exposed to SARS-CoV-2; *A_w_*, amplitude of time-of-week effect in reporting; **φ***_w_*, phase of time-of-week effect in reporting; ρ*_D_*, dispersion parameter of the observation model (NB: this parameter’s definition differs between observation models, so this parameter is only comparable across countries). The y-axis values are log_10_-transformed for every parameter.

In contrast, the choice of the observation model affected the estimates of the basic reproduction number in some countries. Compared with the negative binomial model, the Poisson and binomial models—the two least supported models in each country—produced higher estimates in Belgium (5.7 vs. 5.1, Table S1) but lower estimates in Finland (4.0–4.4 vs. 6.8, Table S2). The estimates were more consistent in Germany and the UK (Tables S3, S6). In all countries, the estimates from the Poisson and binomial models had narrower credible intervals than those from the other observation models. This was particularly notable in Finland, where the best model’s estimates were less certain (99% CI: 4.9–8.6) compared to the Poisson (99% CI: 3.9–4.8) and binomial (99% CI: 3.6–4.2) models. These differences also affected estimates of the effective reproduction number, a key metric for situational awareness and for evaluating epidemic control (Fig. S9).

### 3.4 Sensitivity analyses

We conducted two sensitivity analyses. First, to assess the impact of demographic stochasticity on our results, we implemented a stochastic variant of the process model using a binomial modification of the tau-leap algorithm [29]. We then assessed the log-likelihood of this variant at the MAP estimates using the particle filter algorithm [57]. As shown in Fig. S10, the stochastic and deterministic model variants had similar log-likelihoods in each country, with a maximum absolute difference of about 2 units.

Second, we tested another observation model defined as a discretized normal distribution with mean and variance equal to those of the negative binomial. The negative binomial model was still preferred in every country (Table S7), although the log-likelihood difference was small (at most 4 units). Hence, these results suggest that the choice of the observation models matters beyond the first two moments (mean and variance) of the underlying distribution, although the effect of higher moments was small.

## 4 Discussion

The main goal of this study was to compare various observation models and their impact on the estimated parameters of transmission models of emerging diseases. Using the first wave of COVID-19 as a case study, we applied Bayesian estimation to compare the performance of eight observation models across six countries. Overall, we found large variations in the log-likelihoods of these models across countries, spanning ∼3 orders of magnitude between the best and worst models. The binomial, Poisson, and normal models received little support due to their rigid variance structures. The remaining five models performed considerably better, though their rankings varied across countries. More broadly, our findings emphasize the importance of using flexible observation models to address data imperfections and process model misspecification.

Observation models have received limited attention in the infectious disease modeling literature. Earlier research highlighted the importance of disaggregating incidence data [31] or specifying realistic distributions of case-reporting delays [12,64] for accurate statistical inference of model parameters. However, to our knowledge, only two studies have focused on the selection of the observation model itself. In a study using data simulated from an SIR model and comparing the Poisson, binomial, negative binomial, and beta-binomial (for both the process and observation models), Li et al. reported that distributions that did not account for overdispersion—i.e., Poisson and binomial—lacked flexibility and tended to produce overconfident estimates [22]. In a modeling study based on real-world COVID-19 data in the canton of Geneva, Switzerland, Bouman et al. found that, compared to Poisson and negative binomial observation models, a quasi-Poisson distribution better described the variability in case incidence data (though no formal comparison of the three models was reported) [15]. Although we did not test the quasi-Poisson distribution, its linear mean-variance structure is similar to that of our Student’s *t* distribution, which performed well in two countries (but not in Switzerland).

Hence, our findings are broadly consistent with these earlier studies and emphasize that the choice of observation models is significant, as different models yield different assessments of statistical and predictive uncertainties. Overall, we find that more flexible observation models—those defined by a free dispersion parameter that links the variance to a function of the mean—are more effective at capturing these uncertainties and handling model misspecification.

Although our study focused on SARS-CoV-2, several findings may have broader applicability and serve as a foundation for general recommendations on observation models. First, models like the Poisson and binomial that lack a free dispersion parameter should generally be replaced with more flexible alternatives. In our analyses, these two models consistently performed poorly and underestimated both predictive and parametric uncertainty, especially in key parameters such as the basic reproduction number. The overconfidence of Poisson models—i.e., unrealistically low standard errors—has been noted before for transmission models [22,65] and is generally expected for overdispersed data. This issue is especially problematic during an emerging epidemic, when a reliable assessment of all sources of uncertainty is crucial for epidemic control and prediction.

Compared to the Poisson and binomial models, the normal model had a higher likelihood and produced greater parametric uncertainty. However, it was outperformed by the best models and received no statistical support in any country. Despite having a free dispersion parameter, the model’s fixed variance prevented it from accurately capturing high values of the observed case counts. This is noteworthy because this model is equivalent to nonlinear least squares, a standard method used in nonlinear regression. Emphatically, however, this model’s lack of performance resulted from its constant variance rather than its normal distribution: indeed, the normal model with a more flexible mean-variance structure performed much better.

The performance of the remaining five models was generally comparable, though the negative binomial, overdispersed binomial, and beta-binomial were more consistently ranked among the best. In contrast, the log-normal and overdispersed *t* models performed worse in a few countries. We note that these results apply only to the specific models and data considered here and cannot be generalized to other settings. In addition, despite differences in their log-likelihoods, these five models yielded relatively consistent parameter estimates and comparable parametric and predictive uncertainties. In practice, we recommend testing 2–3 observation models with flexible mean-variance structures, and we caution against less flexible models such as Poisson, binomial, and normal.

Although our study focused on the choice of observation models, our results regarding the process and observation models’ parameters broadly align with those of earlier modeling studies during the first wave of COVID-19 in Europe and the UK. These include a high basic reproduction number and a marked impact of NPIs on transmission, which caused the effective reproduction number to fall below 1 during early spring 2020, before rebounding after the gradual relaxation of control measures in late spring [4,5,32–34,47,66]. Our estimate of the average reporting delay in Germany (∼14 days) is also consistent with previous studies (∼11 days in [5], ∼18 days in [11]). However, we emphasize the different assumptions regarding the delay distribution (Geometric in our study, fixed in [5], and Polya-Aeppli in [11]). Finally, our estimates of the time-of-week effect in case reporting are intuitively reasonable and consistent with other studies that estimated it [5,47].

We acknowledge several limitations of our study. First, all the observation models were based on reported case counts, which are generally the most common endpoints available for infectious diseases. Yet other endpoints—such as counts of hospitalizations or deaths—have been used for statistical inference in SARS-CoV-2 transmission models [4,32,35]. However, as these alternative endpoints may be viewed as scaled-down, delayed versions of reported cases [11], we expect our results to be robust to endpoint definition. Second, we did not incorporate additional sources of variability in the observation model, such as temporal trends in testing capacity, which were pronounced at the start of the COVID-19 pandemic [67]. Relatedly, third, we focused on the first wave of COVID-19 to highlight the importance of observation models when dealing with limited and potentially unstable data during an emerging epidemic. Nevertheless, it is important to note that the performance of these observation models may vary when applied to data from more stable surveillance systems. Fourth, we considered only observation models with at most one free dispersion parameter that controls the variance. However, more complex models with additional parameters could be considered, such as the beta negative binomial distribution (an extension of the negative binomial distribution in which the probability parameter follows a beta distribution).

Another important limitation is that we focused only on transmission models that included a deterministic process model and a stochastic observation model, so all variability in the predictions was due solely to observation noise. This choice was based on the fact that, with few exceptions [1,2,28,30,35], most COVID-19 transmission modeling studies used such partially stochastic models, possibly because statistical inference is quicker and easier with these models than with fully stochastic models, in which both the process and observation models are stochastic. In our study, the likelihoods of both model types at the MAP estimates were similar, suggesting that—at least for the model structure, parameters, and population sizes considered here—the effects of demographic stochasticity were negligible. Importantly, we recognize that stochastic process models are more realistic and flexible and should generally be preferred over their deterministic counterparts [19,31]. Notably, a stochastic process model may alter the inference about the observation model: Since both process and observation noise compete to explain data variability in fully stochastic models, the estimated variance of the observation model may be lower than in partially stochastic models (see Ref. [37] for an empirical illustration based on a negative binomial observation model).

In conclusion, observation models are an overlooked but important aspect of transmission models for infectious diseases. Our results indicate that different observation models can lead to varying levels of predictive and parametric uncertainty, and that poor choices can affect the accuracy and precision of statistical inference for key parameters, such as the basic reproduction number. Therefore, our study could help guide the selection of observation models in future research and improve the design of transmission models that appropriately account for all sources of uncertainty.

## Data Availability

All data and programming code will be permanently archived on GitHub and Zenodo upon publication.

## 6 Paper information

## 6.1 Acknowledgements

We thank Aaron King for his valuable comments and the Infectious Disease Epidemiology lab for a group discussion of the manuscript during a lab meeting. We acknowledge the use of the AI tool PositAI (Version 0.4.8, https://assistant.posit.co/) to assist with code refactoring and documentation; all code logic and final implementations were designed, verified, and approved by the authors. This study was funded by the Max Planck Society.

## 6.2 Author’s Contributions Statement

M.D.d.C conceived of the study design, conducted the analyses, and wrote a first draft of the manuscript. S.C.K. checked the model codes for reproducibility. All authors helped draft and approved the final version of the manuscript.

