## Supplementary Materials for "The impact of observation models on epidemic inference for emerging infectious diseases: SARS-CoV-2 as a case study"

Matthieu Domenech de Cellès[1,*] & Sarah C. Kramer[1]

1. Max Planck Institute for Infection Biology, Infectious Disease Epidemiology group, Berlin, Germany

Corresponding author: Dr. Matthieu Domenech de Cellès. Address: Max Planck Institute for Infection Biology, Charitéplatz 1, Campus Charité Mitte, 10117 Berlin, Germany.

### 1 Supplementary Figures

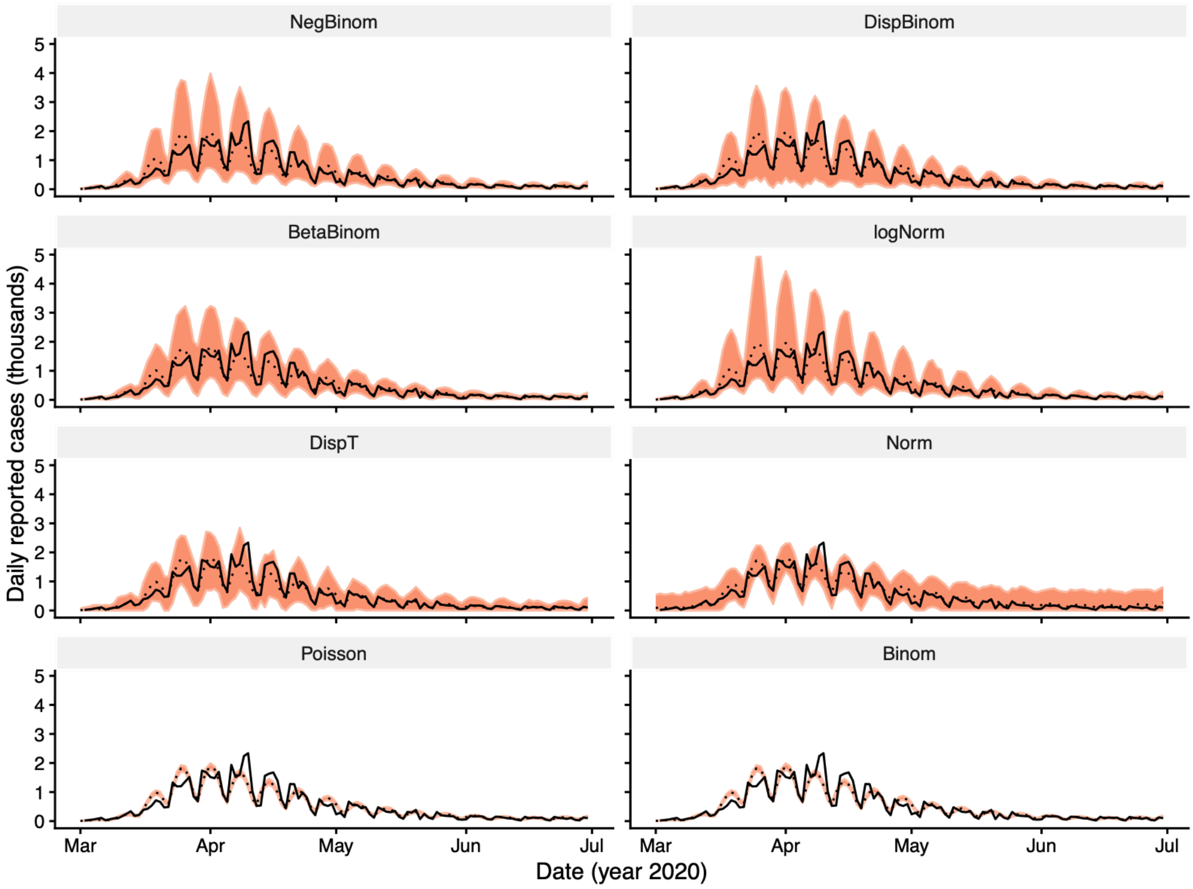


**Figure S1: Models fit in Belgium**. The dotted line (ribbon) shows the mean (99% prediction interval) from 1,000 draws of the observation model, with the process model simulated at the maximum a posteriori estimate of the MCMC chain. Therefore, the predictive variability is only due to the observation model. The solid black line represents the data.

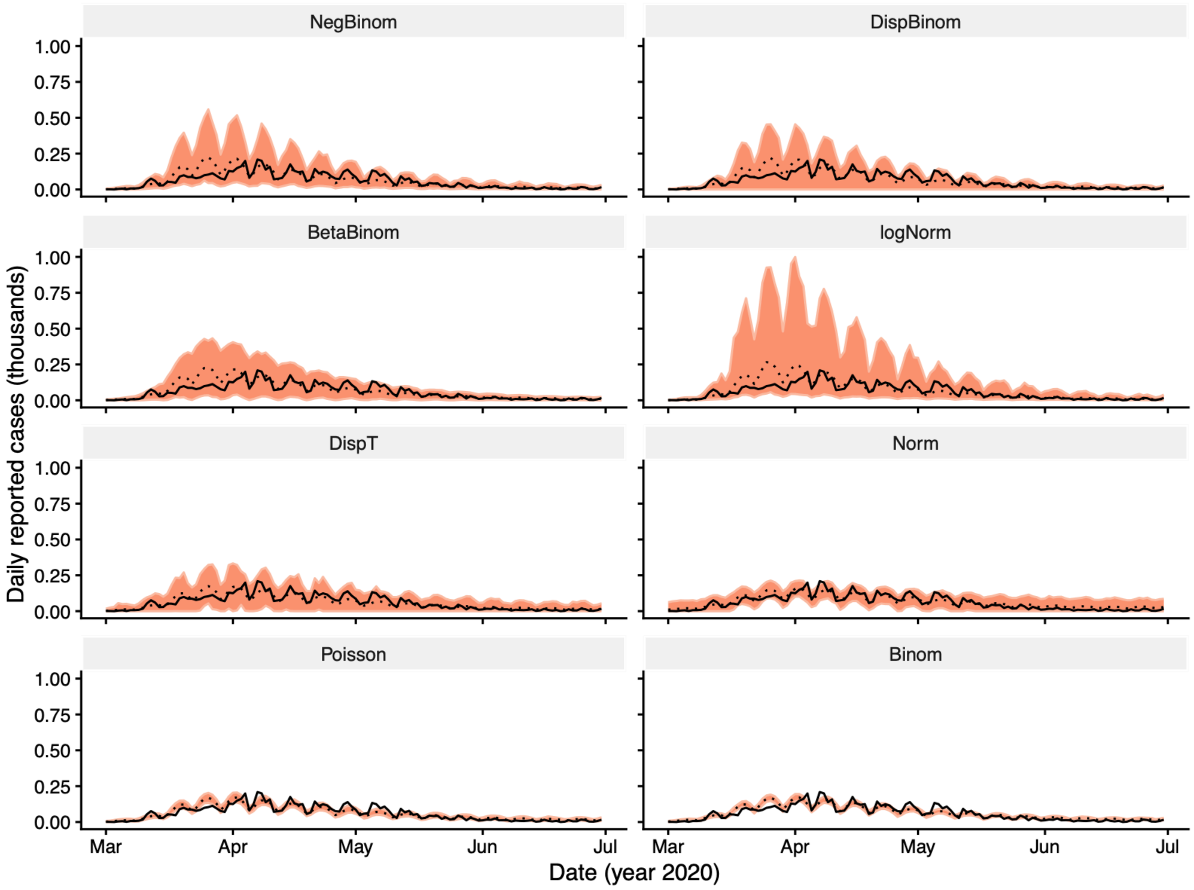


**Figure S2: Models fit in Finland**. The dotted line (ribbon) shows the mean (99% prediction interval) from 1,000 draws of the observation model, with the process model simulated at the maximum a posteriori estimate of the MCMC chain. Therefore, the predictive variability is only due to the observation model. The solid black line represents the data.

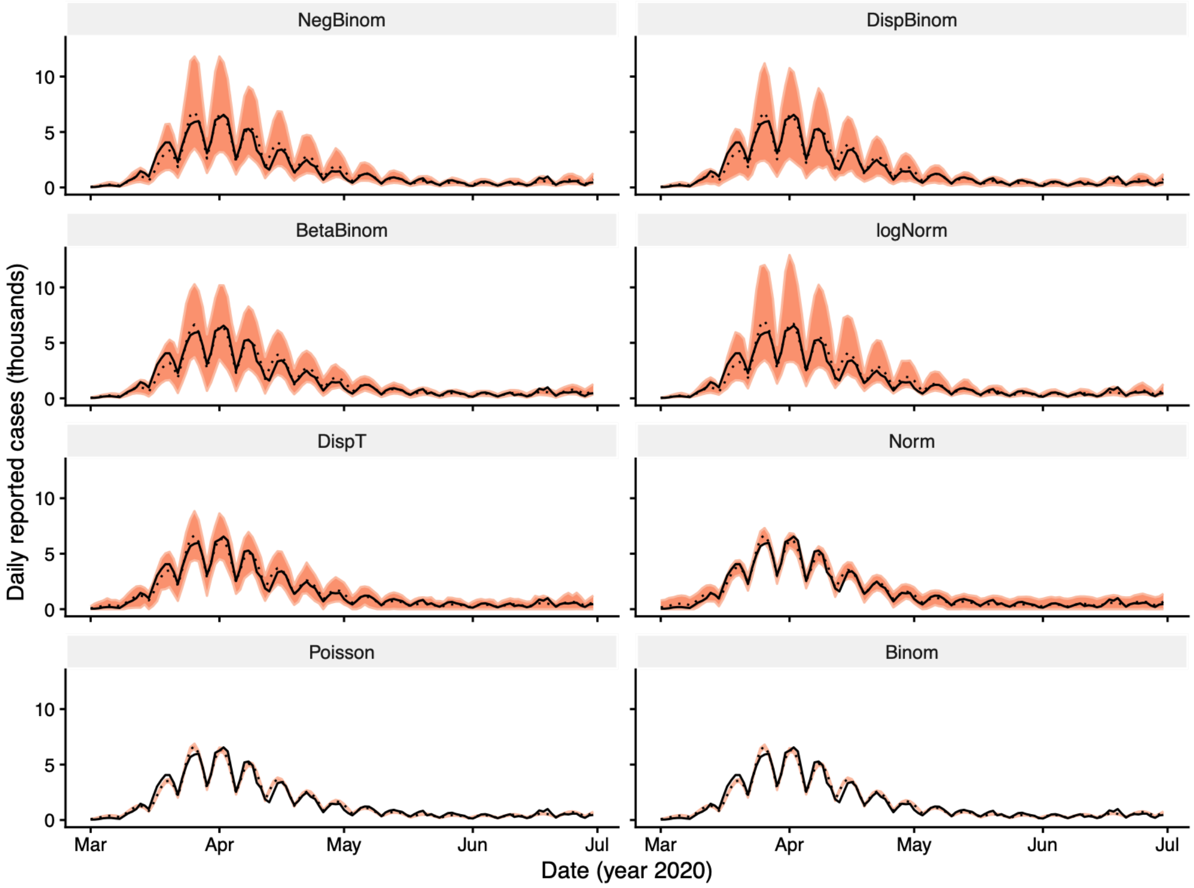


**Figure S3: Models fit in Germany**. The dotted line (ribbon) shows the mean (99% prediction interval) from 1,000 draws of the observation model, with the process model simulated at the maximum a posteriori estimate of the MCMC chain. Therefore, the predictive variability is only due to the observation model. The solid black line represents the data.

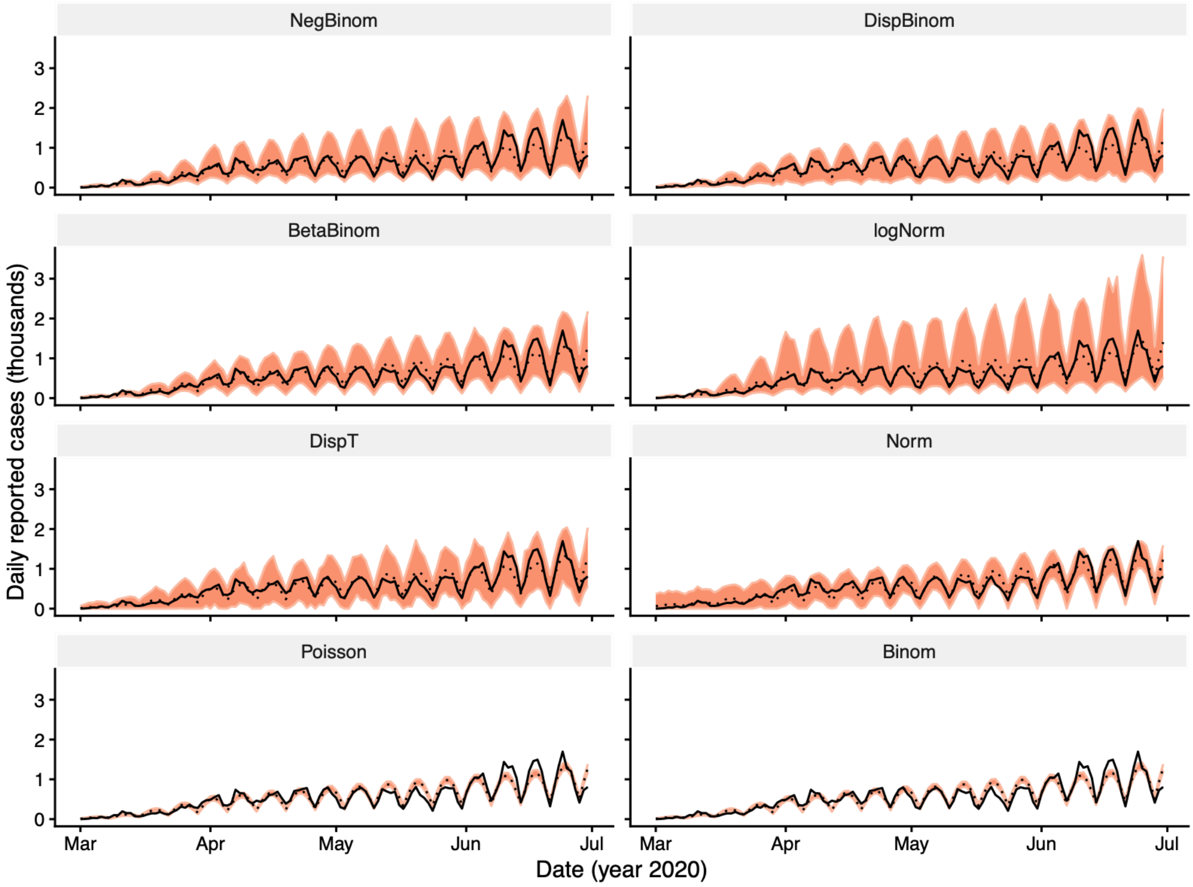


**Figure S4: Models fit in Sweden**. The dotted line (ribbon) shows the mean (99% prediction interval) from 1,000 draws of the observation model, with the process model simulated at the maximum a posteriori estimate of the MCMC chain. Therefore, the predictive variability is only due to the observation model. The solid black line represents the data.

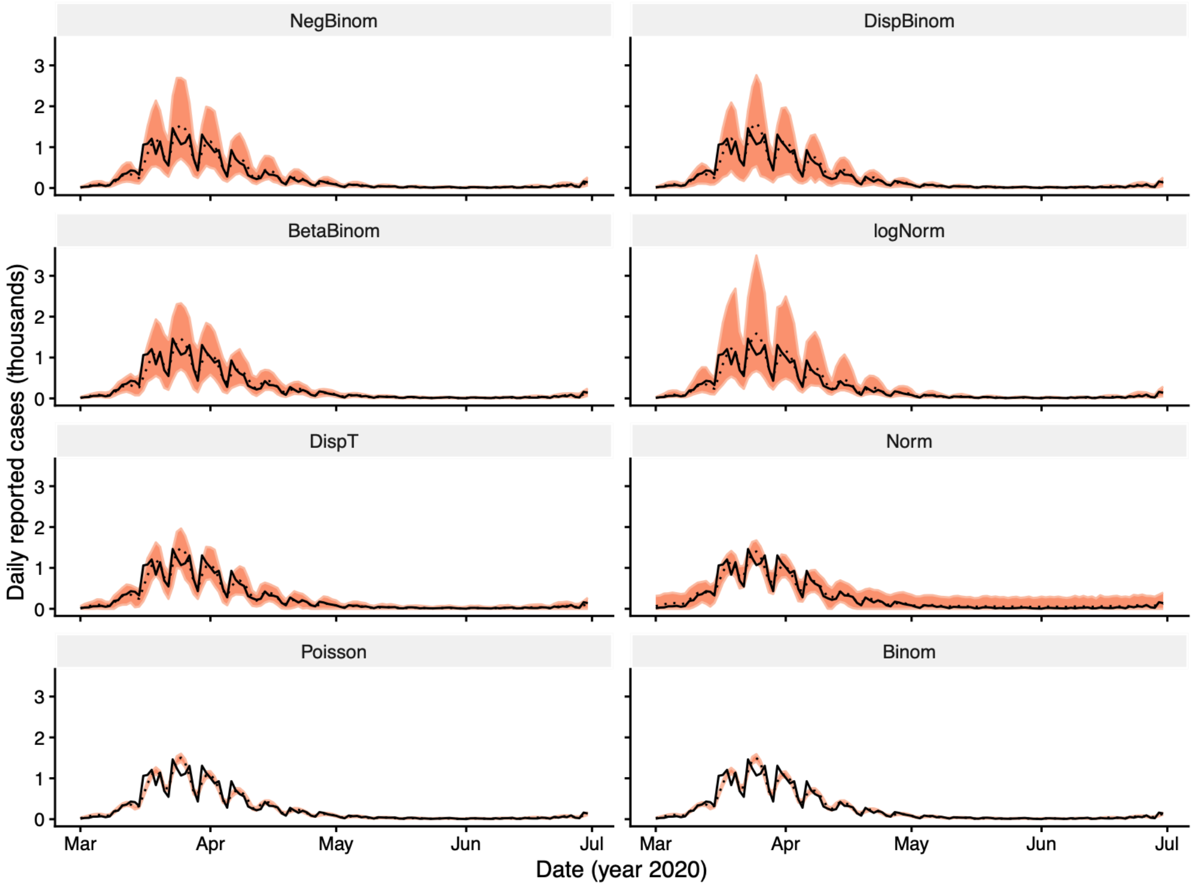


**Figure S5: Models fit in Switzerland**. The dotted line (ribbon) shows the mean (99% prediction interval) from 1,000 draws of the observation model, with the process model simulated at the maximum a posteriori estimate of the MCMC chain. Therefore, the predictive variability is only due to the observation model. The solid black line represents the data.

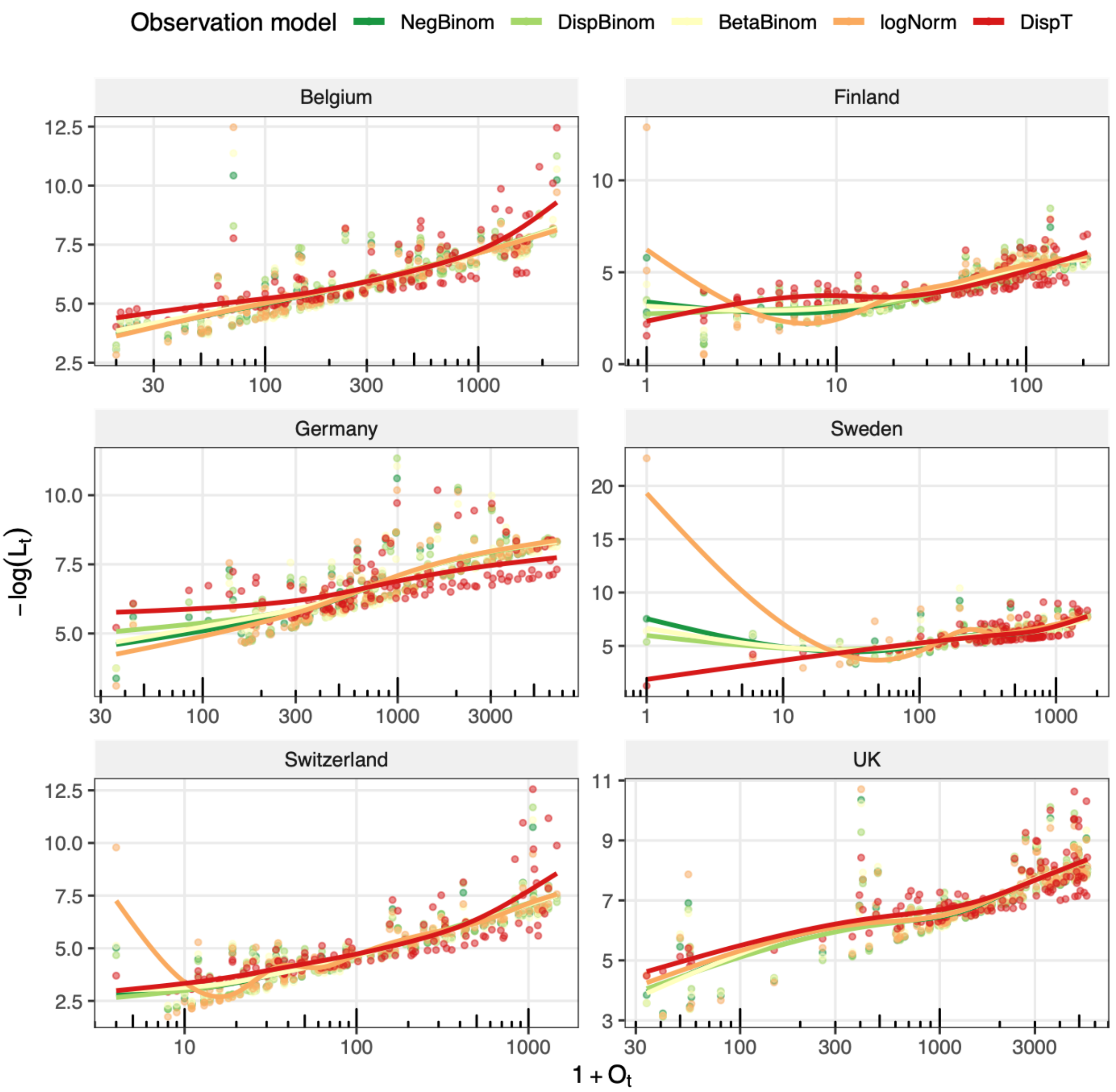


**Figure S6: Comparison of negative pointwise log-likelihoods across the five overdispersed observation models.** For each model, the log-likelihood was computed at the maximum a posteriori (MAP) parameter estimates. The x-axis, representing the observed case counts, is $\log_{10}$-transformed to better visualize the low values. The smooth lines represent fits from generalized additive models.

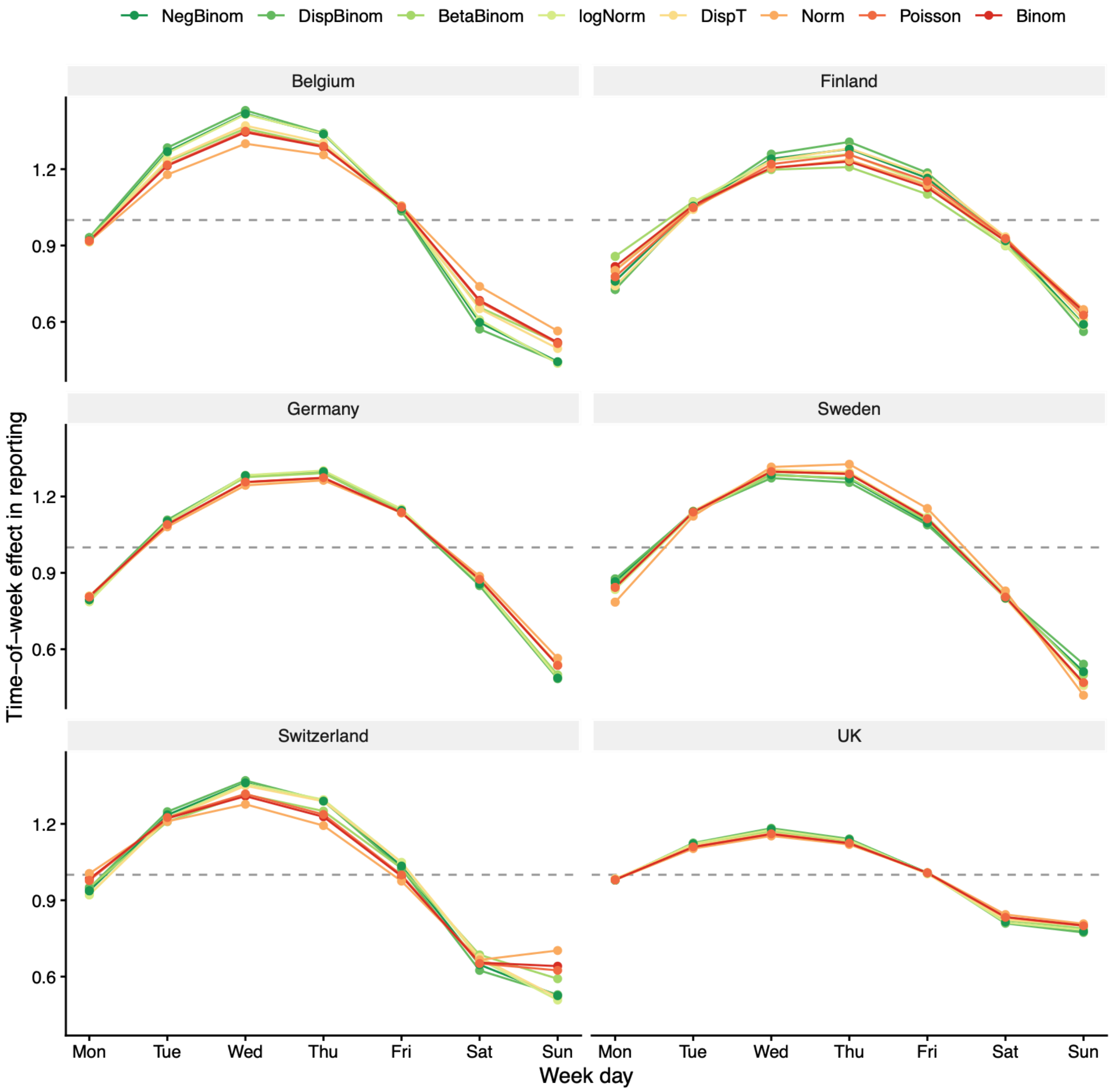


Figure S7: Estimated time-of-week effect in case reporting. For each observation model and country, the parameters were fixed at the maximum a posteriori (MAP) estimates from MCMC.

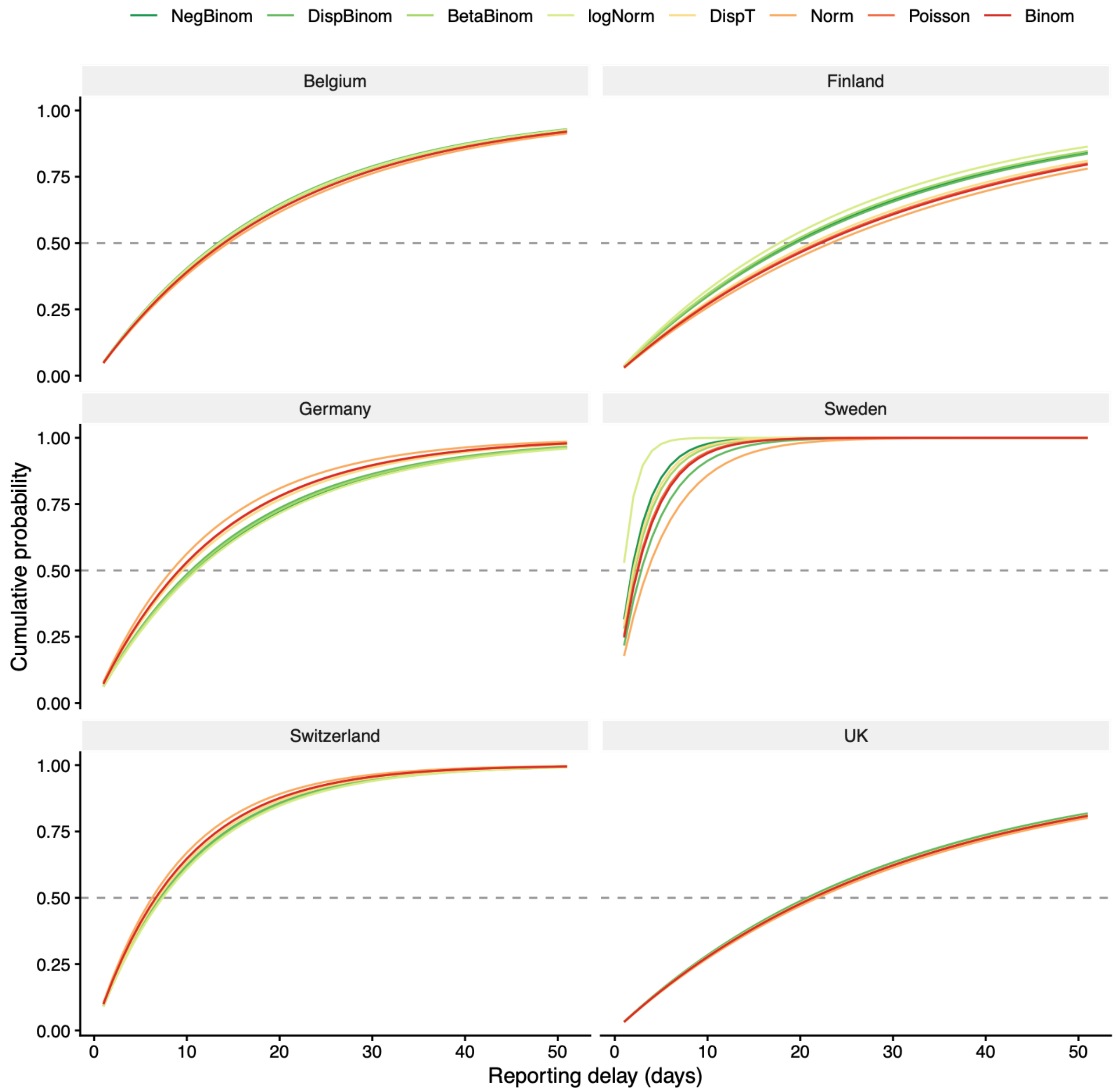


**Figure S8: Estimated case reporting delay.** For each observation model and country, the parameters were fixed at the maximum a posteriori (MAP) estimates from MCMC.

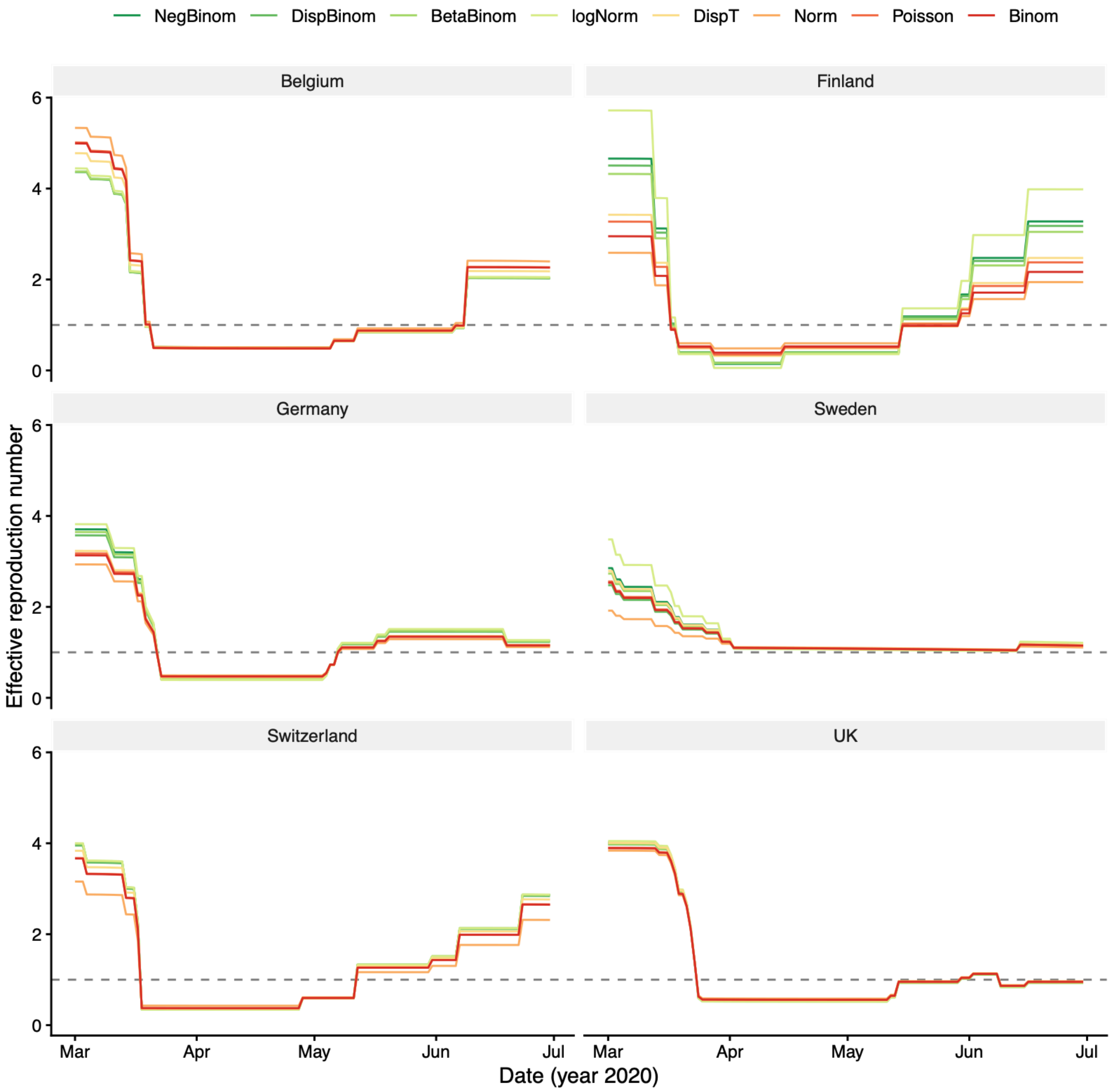


**Figure S9: Estimated effective reproductive number.** For each observation model and country, the parameters were fixed at the maximum a posteriori (MAP) MCMC estimates.

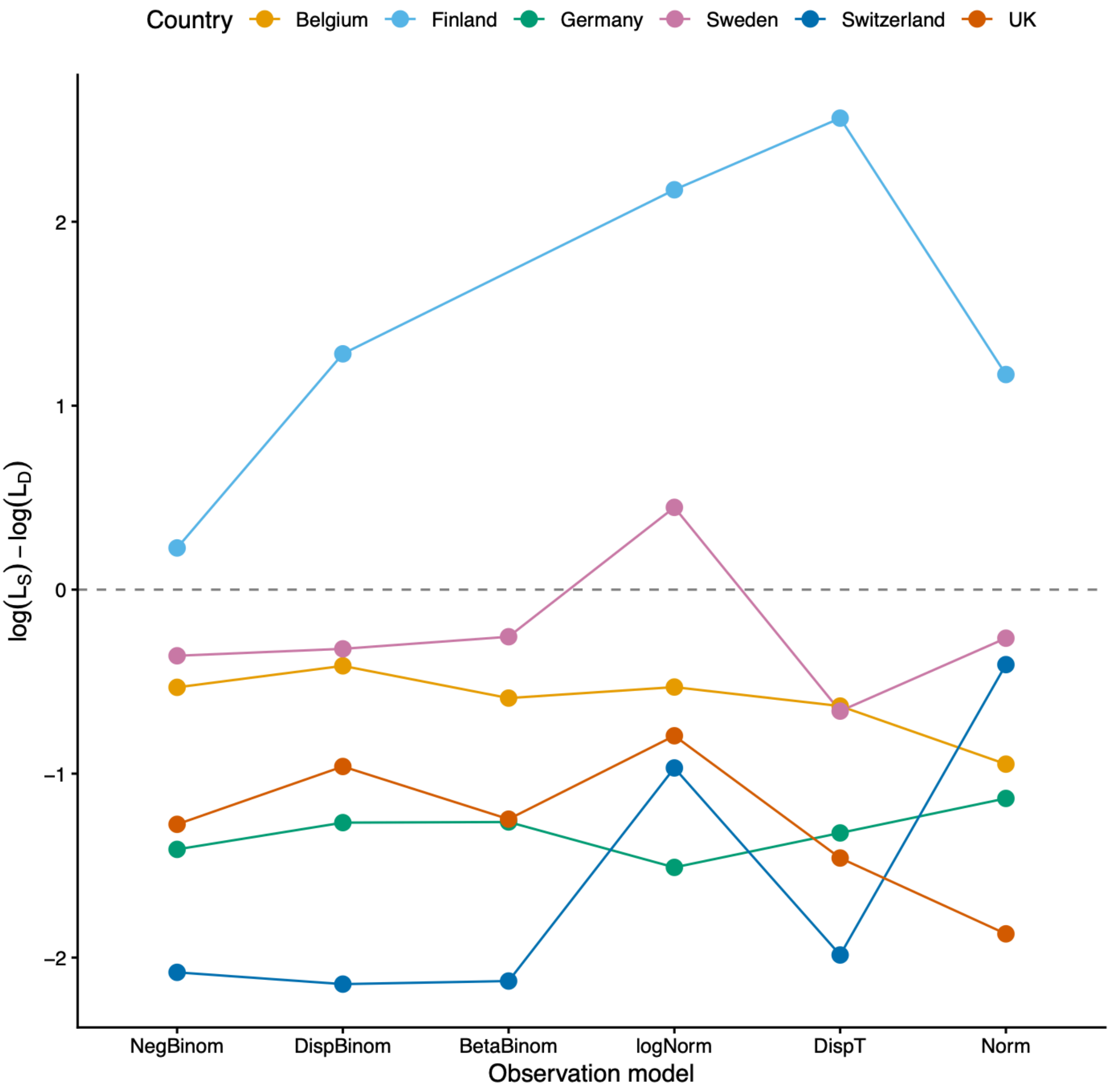


**Figure S10: Log-likelihood difference between the stochastic and deterministic process models.** The likelihood values for both the deterministic ($\log L_D$) and stochastic ($\log L_S$) models were calculated at the maximum a posteriori MCMC estimates. For the stochastic variant, the log-likelihood was estimated as the mean across 10 replicate particle filters, each with 2,000 particles; the corresponding standard errors were typically negligible (<0.1) and are not displayed.

### 2 Supplementary Tables

| Observation Model | $R_0$ | $b_\theta$ | $10^2 \times r_Q$ (day$^{-1}$) | $10^6 \times e_1(0)$ | $A_w$ | $\phi_w$ | $\rho_D$ | $l_{max}$ | DIC | WAIC (SE) | $p_{WAIC}$ |
|---|---|---|---|---|---|---|---|---|---|---|---|
| NegBinom | 5.0 (4.2–5.6) | 1.10 (1.06–1.12) | 5.1 (4.4–5.9) | 34 (21–66) | 0.74 (0.58–0.85) | 0.45 (0.43–0.48) | 0.10 (0.08–0.16) | –718.1 | 1451.8 | 1450.6 (32.0) | 6.4 |
| logNorm | 5.1 (4.3–5.6) | 1.10 (1.06–1.12) | 5.1 (4.3–5.9) | 32 (21–66) | 0.73 (0.59–0.85) | 0.45 (0.43–0.48) | 0.34 (0.30–0.42) | –719.7 | 1454.7 | 1454.2 (33.7) | 6.7 |
| BetaBinom | 5.0 (4.2–5.6) | 1.10 (1.06–1.12) | 5.1 (4.2–6.0) | 33 (21–70) | 0.63 (0.47–0.77) | 0.45 (0.42–0.49) | 0.012 (0.009–0.18) | –721.2 | 1459.6 | 1457.5 (32.5) | 6.6 |
| DispBinom | 5.0 (4.4–5.5) | 1.10 (1.07–1.12) | 5.2 (4.4–6.0) | 35 (22–60) | 0.76 (0.60–0.86) | 0.45 (0.43–0.47) | 0.10 (0.08–0.17) | –722.3 | 1459.0 | 1460.4 (32.0) | 7.8 |
| DispT | 5.5 (4.5–6.7) | 1.10 (1.07–1.15) | 5.0 (4.1–5.8) | 22 (9.0–51) | 0.65 (0.51–0.78) | 0.45 (0.43–0.49) | 0.05 (0.04–0.07) | –743.7 | 1503.5 | 1503.0 (33.5) | 7.9 |
| Norm | 6.1 (4.0–7.0) | 1.10 (1.03–1.15) | 4.8 (3.8–6.2) | 13 (6.7–91) | 0.53 (0.36–0.71) | 0.46 (0.43–0.50) | 0.21 (0.19–0.26) | –819.1 | 1655.8 | 1658.8 (37.1) | 10.7 |
| Poisson | 5.7 (5.5–5.9) | 1.12 (1.11–1.12) | 5.0 (4.8–5.1) | 18 (15–21) | 0.62 (0.60–0.64) | 0.46 (0.45–0.46) | — | –3410.1 | 6832.2 | 7126.9 (1128.1) | 271.8 |
| Binom | 5.7 (5.5–5.9) | 1.12 (1.11–1.12) | 5.0 (4.8–5.1) | 18 (15–21) | 0.61 (0.59–0.63) | 0.46 (0.45–0.46) | — | –3766.5 | 7546.3 | 7891.5 (1278.1) | 312.8 |

**Table S1: MCMC parameter estimates in Belgium.** The values represent the maximum a posteriori (MAP) estimates (99% credible intervals) from 10,000 draws of the posterior distribution. The log-likelihood at the MAP estimates ($l_{max}$) is also shown. DIC: Deviance Information Criterion; WAIC: Widely Applicable Information Criterion; SE: Standard Error; $p_{WAIC}$: effective number of parameters.

| Observation Model | $R_0$ | $b_\theta$ | $10^2 \times r_Q$ (day$^{-1}$) | $10^6 \times e_1(0)$ | $A_w$ | $\phi_w$ | $\rho_D$ | $l_{max}$ | DIC | WAIC (SE) | $p_{WAIC}$ |
|---|---|---|---|---|---|---|---|---|---|---|---|
| NegBinom | 6.4 (4.6–7.9) | 1.40 (1.31–1.42) | 3.6 (3.1–4.1) | 19 (10–42) | 0.50 (0.26–0.67) | 0.54 (0.45–0.59) | 0.18 (0.13–0.31) | –515.2 | 1047.4 | 1044.5 (25.9) | 5.8 |
| DispBinom | 6.1 (4.1–7.7) | 1.40 (1.30–1.40) | 3.6 (2.9–4.1) | 20 (11–57) | 0.55 (0.35–0.69) | 0.54 (0.47–0.58) | 0.20 (0.13–0.35) | –519.5 | 1057.6 | 1054.7 (25.9) | 7.1 |
| BetaBinom | 5.9 (4.0–11.0) | 1.36 (1.28–1.90) | 3.7 (2.9–4.2) | 25 (10–63) | 0.38 (0.20–0.62) | 0.51 (0.43–0.59) | 0.15 (0.12–0.24) | –525.7 | 1069.4 | 1066.6 (24.8) | 6.3 |
| DispT | 4.6 (3.0–7.1) | 1.30 (1.19–1.39) | 3.3 (2.7–3.9) | 40 (12–120) | 0.50 (0.31–0.66) | 0.54 (0.47–0.60) | 0.03 (0.02–0.03) | –530.8 | 1079.3 | 1077.5 (22.9) | 7.8 |
| logNorm | 7.8 (5.9–11.0) | 1.40 (1.40–1.50) | 3.9 (3.2–4.4) | 10 (3.9–24) | 0.47 (0.20–0.68) | 0.52 (0.42–0.60) | 0.59 (0.52–0.74) | –532.4 | 1082.4 | 1082.9 (33.9) | 9.4 |
| Norm | 3.4 (2.4–4.3) | 1.22 (1.05–1.28) | 3.0 (2.3–3.8) | 86 (45–190) | 0.42 (0.24–0.61) | 0.53 (0.47–0.59) | 0.02 (0.02–0.03) | –552.6 | 1121.7 | 1120.2 (22.4) | 6.8 |
| Poisson | 4.4 (3.9–4.8) | 1.29 (1.26–1.31) | 3.2 (3.0–3.4) | 47 (36–64) | 0.46 (0.39–0.52) | 0.54 (0.52–0.55) | — | –863.8 | 1739.8 | 1777.7 (123.6) | 41.9 |
| Binom | 3.9 (3.6–4.2) | 1.26 (1.24–1.28) | 3.1 (3.0–3.3) | 65 (53–78) | 0.41 (0.38–0.48) | 0.53 (0.51–0.54) | — | –1235.5 | 2487.4 | 2557.9 (214.7) | 73.9 |

**Table S2: MCMC parameter estimates in Finland.** The values represent the maximum a posteriori (MAP) estimates (99% credible intervals) from 10,000 draws of the posterior distribution. The log-likelihood at the MAP estimates ($l_{max}$) is also shown. DIC: Deviance Information Criterion; WAIC: Widely Applicable Information Criterion; SE: Standard Error; $p_{WAIC}$: effective number of parameters.

| Observation Model | $R_0$ | $b_\theta$ | $10^2 \times r_Q$ (day$^{-1}$) | $10^6 \times e_1(0)$ | $A_w$ | $\phi_w$ | $\rho_D$ | $l_{max}$ | DIC | WAIC (SE) | $p_{WAIC}$ |
|---|---|---|---|---|---|---|---|---|---|---|---|
| DispT | 4.6 (3.9–5.1) | 1.17 (1.13–1.19) | 7.3 (6.6–8.3) | 2.6 (1.4–6.1) | 0.49 (0.42–0.57) | 0.51 (0.49–0.54) | 0.06 (0.05–0.07) | –830.3 | 1675.5 | 1677.0 (20.9) | 8.8 |
| logNorm | 5.5 (4.9–5.9) | 1.21 (1.19–1.22) | 6.3 (5.8–6.8) | 1.0 (0.7–1.7) | 0.55 (0.45–0.65) | 0.52 (0.48–0.54) | 0.25 (0.22–0.31) | –833.4 | 1682.0 | 1682.5 (27.2) | 7.7 |
| NegBinom | 5.3 (4.7–5.8) | 1.20 (1.18–1.22) | 6.4 (5.9–6.9) | 1.2 (0.71–2.2) | 0.55 (0.45–0.64) | 0.51 (0.48–0.54) | 0.061 (0.048–0.094) | –833.8 | 1683.5 | 1683.9 (26.8) | 8.1 |
| BetaBinom | 5.2 (4.6–5.7) | 1.20 (1.17–1.21) | 6.5 (6.0–7.1) | 1.2 (0.8–2.4) | 0.54 (0.42–0.64) | 0.51 (0.47–0.54) | 0.021 (0.017–0.033) | –838.5 | 1692.9 | 1693.8 (25.9) | 8.5 |
| DispBinom | 5.1 (4.3–5.5) | 1.19 (1.15–1.21) | 6.6 (6.2–7.4) | 1.4 (0.9–3.6) | 0.54 (0.46–0.63) | 0.51 (0.48–0.54) | 0.063 (0.048–0.10) | –837.9 | 1693.2 | 1694.9 (26.3) | 10.2 |
| Norm | 4.1 (3.7–4.5) | 1.14 (1.11–1.17) | 8.3 (7.4–9.2) | 4.5 (2.8–8.0) | 0.48 (0.42–0.55) | 0.52 (0.50–0.54) | 0.27 (0.24–0.34) | –856.7 | 1728.0 | 1732.5 (24.4) | 10.8 |
| Poisson | 4.5 (4.4–4.5) | 1.17 (1.16–1.17) | 7.5 (7.4–7.6) | 2.9 (2.6–3.1) | 0.50 (0.49–0.51) | 0.52 (0.51–0.52) | — | –3587.0 | 7186.6 | 7549.6 (845.2) | 332.2 |
| Binom | 4.4 (4.4–4.5) | 1.16 (1.16–1.16) | 7.6 (7.5–7.7) | 3.1 (2.9–3.3) | 0.50 (0.49–0.51) | 0.51 (0.51–0.52) | — | –4657.2 | 9328.5 | 9819.1 (1124.5) | 447.0 |

**Table S3: MCMC parameter estimates in Germany.** The values represent the maximum a posteriori (MAP) estimates (99% credible intervals) from 10,000 draws of the posterior distribution. The log-likelihood at the MAP estimates ($l_{max}$) is also shown. DIC: Deviance Information Criterion; WAIC: Widely Applicable Information Criterion; SE: Standard Error; $p_{WAIC}$: effective number of parameters.

| Observation Model | $R_0$ | $b_\theta$ | $10^2 \times r_Q$ (day$^{-1}$) | $10^6 \times e_1(0)$ | $A_w$ | $\phi_w$ | $\rho_D$ | $l_{max}$ | DIC | WAIC (SE) | $p_{WAIC}$ |
|---|---|---|---|---|---|---|---|---|---|---|---|
| DispT | 2.9 (2.40–3.6) | 0.96 (0.81–1.07) | 35 (13–440) | 47 (19–120) | 0.57 (0.47–0.67) | 0.49 (0.46–0.53) | 0.04 (0.03–0.06) | –743.8 | 1502.7 | 1503.7 (23.6) | 8.7 |
| BetaBinom | 2.9 (2.5–3.4) | 0.95 (0.87–1.1) | 33 (15–430) | 55 (25–90) | 0.52 (0.42–0.65) | 0.49 (0.46–0.54) | 0.0075 (0.0055–0.012) | –749.9 | 1512.5 | 1515.3 (20.1) | 8.6 |
| NegBinom | 3.0 (2.7–3.5) | 0.98 (0.90–1.07) | 38 (14–470) | 46 (23–87) | 0.52 (0.42–0.63) | 0.49 (0.46–0.53) | 0.07 (0.05–0.11) | –750.9 | 1514.8 | 1518.0 (20.9) | 9.0 |
| DispBinom | 2.6 (2.3–3.2) | 0.89 (0.81–1.00) | 24 (13–390) | 86 (36–130) | 0.50 (0.40–0.61) | 0.48 (0.45–0.53) | 0.07 (0.05–0.12) | –751.9 | 1518.2 | 1520.4 (19.9) | 8.7 |
| Norm | 2.0 (1.4–2.5) | 0.67 (0.30–0.85) | 20 (9.4–390) | 220 (84–670) | 0.61 (0.50–0.71) | 0.51 (0.48–0.53) | 0.13 (0.12–0.16) | –765.9 | 1543.4 | 1546.8 (23.1) | 7.7 |
| logNorm | 3.7 (3.2–4.2) | 1.09 (1.01–1.15) | 75 (23–570) | 19 (10–41) | 0.55 (0.44–0.71) | 0.50 (0.46–0.54) | 0.37 (0.32–0.46) | –781.5 | 1575.9 | 1590.6 (51.5) | 17.9 |
| Poisson | 2.7 (2.6–2.8) | 0.91 (0.87–0.94) | 29 (23–40) | 71 (58–89) | 0.55 (0.53–0.57) | 0.49 (0.49–0.51) | — | –2203.9 | 4420.7 | 4626.1 (467.3) | 223.1 |
| Binom | 2.7 (2.6–2.8) | 0.90 (0.87–0.93) | 28 (23–39) | 75 (60–89) | 0.55 (0.53–0.57) | 0.49 (0.49–0.51) | — | –2389.8 | 4793.3 | 5065.0 (524.8) | 270.8 |

**Table S4: MCMC parameter estimates in Sweden.** The values represent the maximum a posteriori (MAP) estimates (99% credible intervals) from 10,000 draws of the posterior distribution. The log-likelihood at the MAP estimates ($l_{max}$) is also shown. DIC: Deviance Information Criterion; WAIC: Widely Applicable Information Criterion; SE: Standard Error; $p_{WAIC}$: effective number of parameters.

| Observation Model | $R_0$ | $b_\theta$ | $10^2 \times r_Q$ (day$^{-1}$) | $10^6 \times e_1(0)$ | $A_w$ | $\phi_w$ | $\rho_D$ | $l_{max}$ | DIC | WAIC (SE) | $p_{WAIC}$ |
|---|---|---|---|---|---|---|---|---|---|---|---|
| NegBinom | 5.3 (4.7–5.8) | 1.28 (1.25–1.29) | 9.5 (8.9–10.0) | 9.3 (6.1–16.0) | 0.64 (0.49–0.75) | 0.45 (0.42–0.48) | 0.07 (0.05–0.12) | –578.7 | 1173.1 | 1172.4 (34.4) | 7.0 |
| BetaBinom | 5.3 (4.6–5.7) | 1.28 (1.25–1.29) | 9.5 (8.9–10.0) | 9.1 (6.4–16.0) | 0.56 (0.39–0.69) | 0.45 (0.41–0.49) | 0.02 (0.01–0.03) | –580.3 | 1177.0 | 1176.0 (34.0) | 7.0 |
| logNorm | 5.3 (4.7–5.9) | 1.28 (1.25–1.30) | 9.4 (8.7–10.0) | 9.3 (5.9–16.0) | 0.63 (0.48–0.75) | 0.45 (0.43–0.49) | 0.32 (0.29–0.40) | –583.2 | 1181.3 | 1181.6 (36.1) | 7.3 |
| DispBinom | 5.3 (4.7–5.8) | 1.27 (1.25–1.29) | 9.7 (9.1–11.0) | 9.7 (6.3–16.0) | 0.65 (0.48–0.76) | 0.44 (0.42–0.47) | 0.08 (0.05–0.13) | –582.8 | 1180.8 | 1182.2 (35.8) | 8.4 |
| DispT | 5.1 (4.4–5.7) | 1.27 (1.24–1.29) | 10.0 (9.3–11.0) | 11.0 (6.3–20.0) | 0.62 (0.49–0.71) | 0.46 (0.43–0.48) | 0.03 (0.02–0.04) | –608.1 | 1230.8 | 1236.3 (39.2) | 12.1 |
| Norm | 4.1 (3.5–4.7) | 1.22 (1.18–1.29) | 11.0 (7.3–14.0) | 28.0 (16.0–66.0) | 0.49 (0.35–0.61) | 0.42 (0.38–0.46) | 0.10 (0.09–0.13) | –739.1 | 1494.2 | 1506.6 (34.2) | 17.8 |
| Poisson | 4.9 (4.7–5.0) | 1.26 (1.25–1.27) | 10.0 (10.0–11.0) | 14.0 (12.0–16.0) | 0.56 (0.53–0.59) | 0.43 (0.43–0.44) | — | –1429.3 | 2870.4 | 3049.7 (473.0) | 169.1 |
| Binom | 4.9 (4.7–5.0) | 1.26 (1.25–1.27) | 10.0 (10.0–11.0) | 14.0 (12.0–16.0) | 0.54 (0.51–0.57) | 0.43 (0.42–0.44) | — | –1691.5 | 3396.5 | 3641.0 (602.8) | 219.1 |

**Table S5: MCMC parameter estimates in Switzerland.** The values represent the maximum a posteriori (MAP) estimates (99% credible intervals) from 10,000 draws of the posterior distribution. The log-likelihood at the MAP estimates ($l_{max}$) is also shown. DIC: Deviance Information Criterion; WAIC: Widely Applicable Information Criterion; SE: Standard Error; $p_{WAIC}$: effective number of parameters.

| Observation Model | $R_0$ | $b_\theta$ | $10^2 \times r_Q$ (day$^{-1}$) | $10^6 \times e_1(0)$ | $A_w$ | $\phi_w$ | $\rho_D$ | $l_{max}$ | DIC | WAIC (SE) | $p_{WAIC}$ |
|---|---|---|---|---|---|---|---|---|---|---|---|
| NegBinom | 4.6 (4.3–4.9) | 1.11 (1.09–1.14) | 3.3 (2.8–3.8) | 2.2 (1.5–3.3) | 0.31 (0.22–0.40) | 0.44 (0.41–0.48) | 0.03 (0.02–0.05) | –859.0 | 1733.9 | 1732.5 (27.2) | 6.7 |
| DispBinom | 4.5 (4.2–4.8) | 1.11 (1.08–1.13) | 3.3 (2.9–3.8) | 2.4 (1.7–3.6) | 0.32 (0.22–0.40) | 0.44 (0.40–0.48) | 0.03 (0.03–0.05) | –859.6 | 1733.7 | 1733.3 (26.9) | 6.3 |
| BetaBinom | 4.5 (4.2–4.8) | 1.11 (1.09–1.13) | 3.2 (2.9–3.8) | 2.4 (1.7–3.5) | 0.30 (0.20–0.39) | 0.44 (0.40–0.48) | 0.009 (0.007–0.013) | –861.5 | 1737.8 | 1737.1 (27.3) | 6.5 |
| logNorm | 4.6 (4.3–4.9) | 1.11 (1.09–1.14) | 3.3 (2.8–3.7) | 2.1 (1.5–3.1) | 0.31 (0.21–0.40) | 0.44 (0.40–0.49) | 0.19 (0.17–0.24) | –862.1 | 1740.0 | 1739.9 (27.0) | 7.6 |
| DispT | 4.6 (4.0–5.0) | 1.10 (1.07–1.13) | 3.3 (2.9–3.8) | 2.1 (1.2–4.6) | 0.29 (0.19–0.37) | 0.44 (0.40–0.48) | 0.07 (0.05–0.08) | –874.6 | 1764.2 | 1764.1 (24.8) | 7.2 |
| Norm | 4.4 (3.5–5.0) | 1.08 (1.03–1.11) | 3.2 (2.7–3.8) | 2.8 (1.0–9.3) | 0.27 (0.19–0.35) | 0.44 (0.40–0.48) | 0.41 (0.36–0.51) | –906.7 | 1829.0 | 1827.8 (16.9) | 6.3 |
| Poisson | 4.4 (4.4–4.5) | 1.10 (1.09–1.10) | 3.2 (3.2–3.3) | 2.5 (2.3–2.8) | 0.28 (0.27–0.29) | 0.44 (0.44–0.45) | — | –4316.7 | 8645.0 | 8963.8 (974.3) | 299.5 |
| Binom | 4.4 (4.4–4.5) | 1.09 (1.09–1.10) | 3.2 (3.2–3.3) | 2.6 (2.4–2.9) | 0.28 (0.27–0.29) | 0.44 (0.44–0.45) | — | –5285.6 | 10583.8 | 10980.9 (1227.0) | 377.9 |

**Table S6: MCMC parameter estimates in the UK.** The values represent the maximum a posteriori (MAP) estimates (99% credible intervals) from 10,000 draws of the posterior distribution. The log-likelihood at the MAP estimates ($l_{max}$) is also shown. DIC: Deviance Information Criterion; WAIC: Widely Applicable Information Criterion; SE: Standard Error; $p_{WAIC}$: effective number of parameters.

| **Country** | **$l_{max}$ (negative binomial model)** | **$l_{max}$ (matching normal model)** |
|---|---|---|
| Belgium | –718.1 | –722.2 |
| Finland | –515.2 | –519.3 |
| Germany | –833.8 | –837.8 |
| Sweden | –750.9 | –751.5 |
| Switzerland | –578.7 | –582.7 |
| UK | –859.0 | –859.4 |

**Table S7: Comparison of negative binomial and matching normal observation models.** The matching normal model represents a discretized normal distribution whose first two moments (mean and variance) equal those of the negative binomial distribution. The values $l_{max}$ represent the log-likelihood at the MAP estimates ($l_{max}$).
